# TMAO and its precursors in relation to host genetics, gut microbial composition, diet, and clinical outcomes: Meta-analysis of 5 prospective population-based cohorts

**DOI:** 10.1101/2022.09.01.22279510

**Authors:** Sergio Andreu-Sánchez, Shahzad Ahmad, Alexander Kurilshikov, Marian Beekman, Mohsen Ghanbari, Martijn van Faassen, Inge C.L. van den Munckhof, Marinka Steur, Amy Harms, Thomas Hankemeier, M. Arfan Ikram, Maryam Kavousi, Trudy Voortman, Robert Kraaij, Mihai G. Netea, Joost H.W. Rutten, Niels P. Riksen, Alexandra Zhernakova, Folkert Kuipers, P. Eline Slagboom, Cornelia M. van Duijn, Jingyuan Fu, Dina Vojinovic

## Abstract

Trimethylamine N-oxide (TMAO) is a circulating microbiome-derived metabolite implicated in the development of atherosclerosis and cardiovascular disease (CVD). We investigated whether plasma levels of TMAO, its precursors (betaine, carnitine, deoxycarnitine, choline) and TMAO-to-precursor ratios associate with clinical outcomes, including CVD and mortality. This was followed by an in-depth analysis of their genetic, gut microbial and dietary determinants. The analyses were conducted in five Dutch prospective cohort studies including 7,834 individuals. To further investigate association results, Mendelian Randomization (MR) was also explored. We found only plasma choline levels (hazard ratio (HR) 1.17, (95% CI 1.07; 1.28)) and not TMAO to be associated with CVD risk. Our association analyses uncovered 10 genome-wide significant loci, including novel genomic regions for betaine (6p21.1, 6q25.3), choline (2q34, 5q31.1) and deoxycarnitine (10q21.2, 11p14.2) comprising several metabolic gene associations, e.g., *CPS1* or *PEMT*. Furthermore, our analyses uncovered 68 gut microbiota associations, mainly related to TMAO-to-precursors ratios and the *Oscillospiraceae* family and 16 associations of food groups and metabolites including fish-TMAO, meat-carnitine and plant-based food-betaine associations. No significant association was identified by MR approach. Our analyses provide novel insights into the TMAO pathway, its determinants and pathophysiological impact in the general population.

## INTRODUCTION

There is a growing interest in the role of gut microbiome-related metabolites in cardiovascular disease (CVD).^1, 2^ Trimethylamine N-oxide (TMAO) in particular has received a lot of attention as a potential promoter of CVD and atherosclerosis.^1^ Elevated fasting plasma levels of TMAO have been associated with increased risk for CVD and mortality independently of traditional risk factors in clinical studies.^3, 4, 5,6, 7, 8^ However, investigations on TMAO have mainly been conducted in individuals with high risk of CVD, existing disease or multimorbidity while studies on general population are scarce.^9^ Proposed mechanisms through which TMAO may promote the development of atherosclerosis and CVD include vascular inflammation, activation of platelets, disturbance of bile acid metabolism and inhibition of reverse cholesterol transport.^10^ Yet, the actual mode of action in disease development may be context dependent.

Variation in circulating levels of TMAO is driven by a complex interplay of multiple determinants, such as host genetics, gut microbiome, diet and kidney function.^11^ TMAO can be acquired directly from the diet from fish but it is mainly produced by gut microbiota from the dietary precursors choline, L-carnitine, the carnitine-derived metabolite deoxycarnitine (also known as γ-butyrobetaine) and betaine.^12^ Gut microbiota convert these ubiquitous dietary components into trimethylamine (TMA) which is subsequently absorbed from intestine, transported to the liver and oxidized into TMAO by hepatic flavin monooxygenases (FMOs), followed by its distribution to different tissues or kidney clearance.^13^ However, the extent to which consumption of different animal-based foods may affect plasma TMAO levels is debated,^14, 15, 16^ and TMA producers in the human gut microbiome have not been well-characterized. Association studies point to several genera, however, published results are in most cases heterogeneous among studies.^12, 17, 18, 19, 20, 21^ On the other hand, several studies have focused on identifying microbial genomes harboring the metabolic potential to produce TMA.^22, 23^ Even though, *Proteobacteria* and *Firmicutes* phyla had the most predicted TMA-production potential,^23^ reports suggest that there is no association between gene abundance levels and TMAO plasma concentrations.^24^ At the same time, human genetic variation also contributes to TMAO variability. Rare genetic mutations in *FMO* type 3 gene (*FMO3*) have been shown to affect oxidation of TMA to TMAO.^25^ However, the role of common genetic variation in TMAO homeostasis remains to be elucidated. As identifying potential drivers for alterations in circulating TMAO levels could have preventive and therapeutic implications for CVD,^26^ a number of studies have explored determinants of TMAO.^27, 12, 21^ However, these cross-sectional multi-omics studies lack in most cases large sample sizes and are limited to explore TMAO variability while all their precursors are overlooked. The physiological impact of choline and betaine associations with CVD risk remain controversial.^28, 29, 30, 31^ Importantly, the major sources of intraindividual variability of these metabolites, together with carnitine and deoxycarnitine, remain unknown.

In addition, CVD risk has often been regarded as a sex-related disease, with clear prevalence differences between males and females.^32^ No studies to date have investigated whether the effect of TMAO metabolites on CVD risk might be sex-related and whether sex might differentially affect the factors that shape plasma concentrations of these metabolites.

Therefore, our aim was to conduct an in-depth study to examine associations of plasma levels of TMAO and its precursors (betaine, carnitine, deoxycarnitine, choline) with clinical outcomes, including CVD incidence and mortality and to investigate their genetic, gut microbial and dietary determinants in the setting of five Dutch prospective cohort studies mainly based on the general population. Additionally, we also investigated ratios of TMAO to its dietary precursors^33^ as they could provide important information about the TMAO biosynthesis pathway in relation to CVD. Lastly, we aimed to assess the potential causal nature of the relationships between TMAO and its precursors with CVD by using a Mendelian Randomization approach.

## RESULTS

### Characteristics of the study population

An overview of the study design is depicted in **Figure 1**. Our study population included 7,834 participants from five Dutch prospective cohort studies including the Rotterdam Study I-4, Rotterdam Study III-2, Leiden Longevity Study (LLS), LifeLines-DEEP (LLD) and 300-Obese cohort (300-OB). A description of contributing cohorts is provided in **Supplementary Material** and descriptive characteristics of study participants are shown in **Table 1**. The mean age of study participants ranged from 43.4 years (sd = 14.2) in the LLD to 75.1 years (sd = 6.1) in the Rotterdam Study I-4. The sex ratio of participants was roughly balanced, with slightly more females in most of the participating studies (up to 58%), with the exception of 300-OB in which the majority of participants were males (55 %). Detailed information on clinical outcomes such as CVD and mortality, host genetics, gut microbiome composition and diet was available for cohorts.

**Figure 1.**
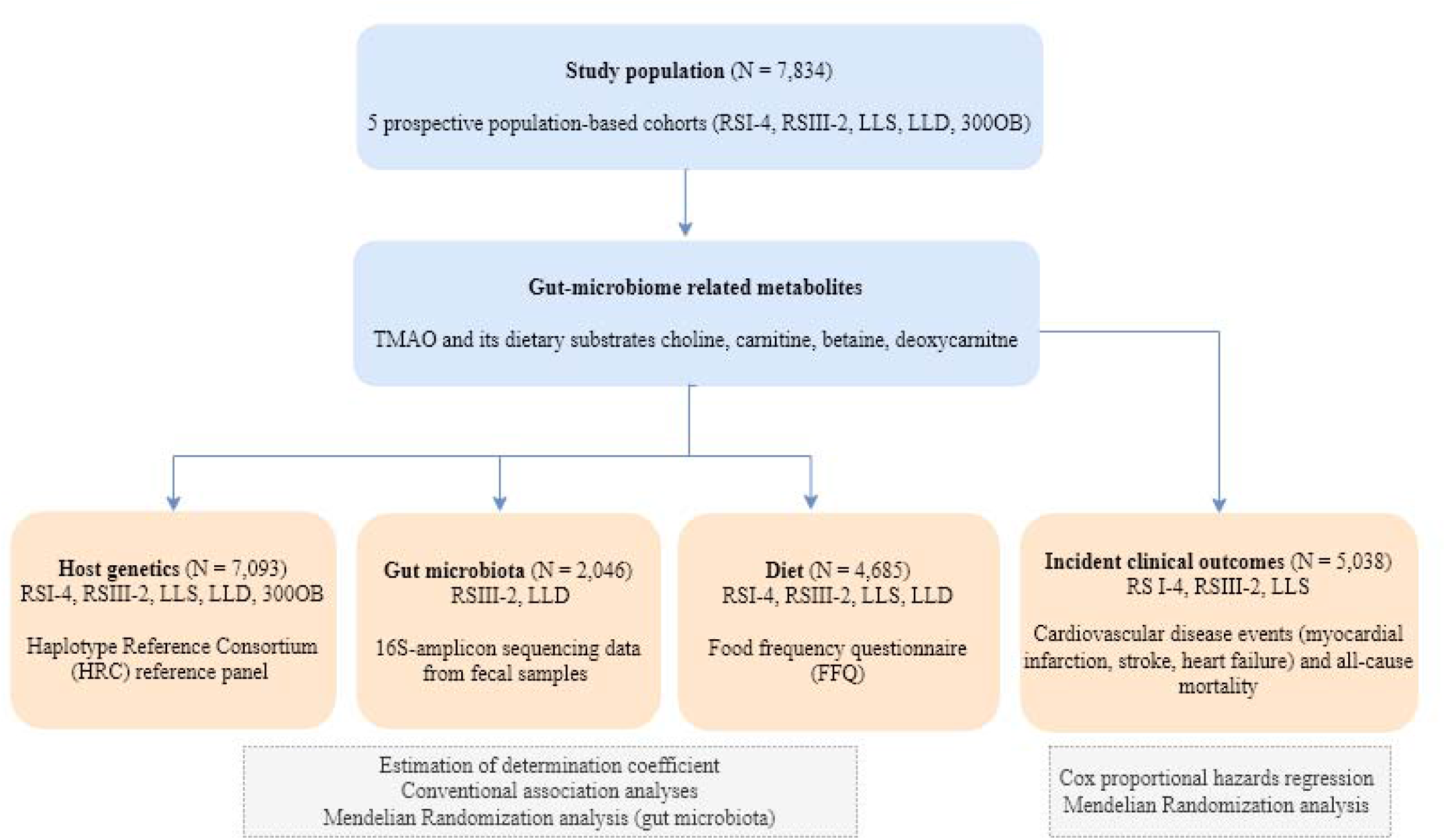
Overview of study design and performed analyses. Study population included participants from Rotterdam Study I-4 (RSI-4), Rotterdam Study III-2 (RSIII-2), Leiden Longevity Study (LLS), LifeLines-DEEP (LLD) and 300-Obese cohort (300-OB).

**Table 1.** Descriptive characteristics of study population.

|  | **Rotterdam**<br>**Study I-4** | **Rotterdam Study**<br>**III-2** | **LLS** | **LLD** | **300-OB** |
| --- | --- | --- | --- | --- | --- |
| N | 2556 | 1377 | 2158 | 1650 | 302 |
| Age (years), mean (sd) | 75.14 (6.08) | 62.68 (5.82) | 59.12 (6.71) | 43.94<br>(14.15) | 67.05 (5.39) |
| Women, N (%) | 1486 (58.14) | 748 (54.32) | 1208 (55.98) | 833 (57.52) | 167 ( 55.29) |
| Smoking, N (%) | 301 (12.05) | 238 (17.32) | 241 (11.17) | 289 (19.63) | 26 (8.63) |
| Diabetes, N (%) | 346 (13.61) | 123 (8.94) | 90 (4.17) | 28 (1.7) | 37 (12.25) |
| Hypertension, N (%) | 2182 (85.60) | 752 (54.77) | 159 (7.37) | 321<br>(19.47%) | 175 (57.94) |
| LDLC (mmol/L), mean (sd) | 1.63 (0.44) | 1.76 (0.51) | 1.60 (0.47) | 3.14 (0.91) | 4.13 (0.96) |
| HDLC (mmol/L), mean (sd) | 1.40 (0.28) | 1.37 (0.35) | 1.49 (0.33) | 1.52 (0.40) | 1.33 (0.31) |
| Serum triglycerides (mmol/L), mean (sd) | 1.38 (0.57) | 1.33 (0.62) | 1.55 (0.82) | 1.16 (0.86) | 1.83 (1.02) |
| Lipid-lowering medication, N (%) | 583 (22.90) | 362 (26.51) | 207 (9.59) | 29 (2.55) | 83 (27.48) |
| BMI (kg/m<sup>2</sup>), mean (sd) | 27.43 (4.13) | 27.44 (4.50) | 25.43 (3.57) | 25.30 (4.22) | 30.73 (3.48) |
| eGFR* | 79.41 (13.10) | 95.31 (9.29) | 94.88 (11.60) | 73.13(12.6) | 80.4 (15.68) |
| Incident cardiovascular disease | 544 | 9 | 85 | 23 (1.5%) | - |
| Follow-up time cardiovascular disease,<br>mean (years) | 8.62 (3.49) | 2.48 | 10.7 (1.77) | - | - |
| Incident mortality events | 1295 | 37 | 209 | - | - |
| Follow-up time mortality, mean (years) | 10.15 (3.88) | 2.48 (1.17) | 12.74 (2.22) | - | - |

Abbreviations: LLS - Leiden Longevity Study, LLD - LifeLines-DEEP, 300-OB - 300-Obese cohort, LDLC - cholesterol in low-density lipoproteins, HDLC - high-density lipoproteins, BMI – body-mass index, eGFR – estimated glomerular filtration rate;
\*eGFR – calculated using the sing the Chronic Kidney Disease Epidemiology Collaboration equation (CKD-EPI).

### Characteristics of gut microbiome related metabolites

Plasma levels of TMAO and its precursors betaine, carnitine, deoxycarnitine and choline were measured in all participating cohorts. Descriptive characteristics of TMAO and its precursors betaine, carnitine, deoxycarnitine and choline and the methods used to quantify their plasma concentrations are shown in **Supplementary Table 1**. Phenotypic correlations between metabolites and their ratios are displayed in **Supplementary Figure 1**. Weak phenotypic correlations were observed between TMAO and its precursors (r^2^ ≤ 0.20) and moderate correlations between the TMAO precursors (0.30 ≤ r^2^ ≤ 0.44).

### Incident clinical outcomes are associated with TMAO precursors but not with TMAO

To assess the relationship between metabolites and incident clinical outcomes including CVD and mortality, Cox proportional hazards regression models with age as time scale were used. In total, 571 incident CVD events and 1,440 mortality events were observed among up to 5,011 participants from Rotterdam Study I-4, Rotterdam Study III-2 and LLS. The results of association analyses are displayed in **Supplementary Tables 2** and **3**. A statistically significant association was observed between higher levels of choline and risk of CVD (hazard ratio (HR) 1.17, (95% CI 1.07; 1.28)) after adjusting for sex, body-mass index (BMI), hypertension, diabetes, cholesterol in low-density lipoproteins (LDLC) and high-density lipoproteins (HDLC), serum triglycerides (TG), use of lipid-lowering medication, current smoking, fasting status (if appropriate) (model 1) and multiple testing. The association did not change after further adjustment for estimated glomerular filtration rate (eGFR) (model 2). The direction of effect size was concordant among the cohorts and no differences were observed between males and females (**Supplementary Table 2**). Additionally, a nominal significant association was observed between betaine and risk of CVD (HR 1.10; 95% CI 1.00; 1.21). This association was driven by a strong association in males (HR 1.23; 95% CI 1.07; 1.43). TMAO was not associated neither with CVD risk or mortality. Analyses were repeated using quartiles of TMAO and also here no association was observed with clinical outcomes.

To assess potential causal effect, we performed a MR analysis. The results did not provide evidence for any causal effect of TMAO or its precursor metabolites on CVD (**Supplementary Table 4**).

### Drivers of variation in gut microbiome related metabolite levels

We next investigated the sources of variability of each of the measured metabolites. The combined effect of host genetics, gut microbiome and dietary variation in explaining the variability of plasma metabolite levels and ratios was evaluated by fitting linear regularized additive models (elastic net) on a train set and by estimating the determination coefficient (R^2^) on a test set (see details in **Method**s). We trained two models, using LLD or Rotterdam Study III-2 as the train set and the left-out cohort as the test set, respectively (**Figure 2**). Genetic contributions to metabolite variability was large for TMAO precursors but small for TMAO and TMAO-to-precursor ratios, although it showed low replicability in the testing cohort. We observed similar pattern regarding microbial features. For both LLD and Rotterdam Study III-2 trained models, a large proportion of TMAO and TMAO ratios variability could be explained by the microbiome in the training set. However, this effect was lost in the test sets. On the other hand, diet showed small but consistent effects between cohorts, while anthropometrics effects seemed to be larger in LLD, both in the Rotterdam Study III-2 -trained and LLD-trained models.

**Figure 2.**
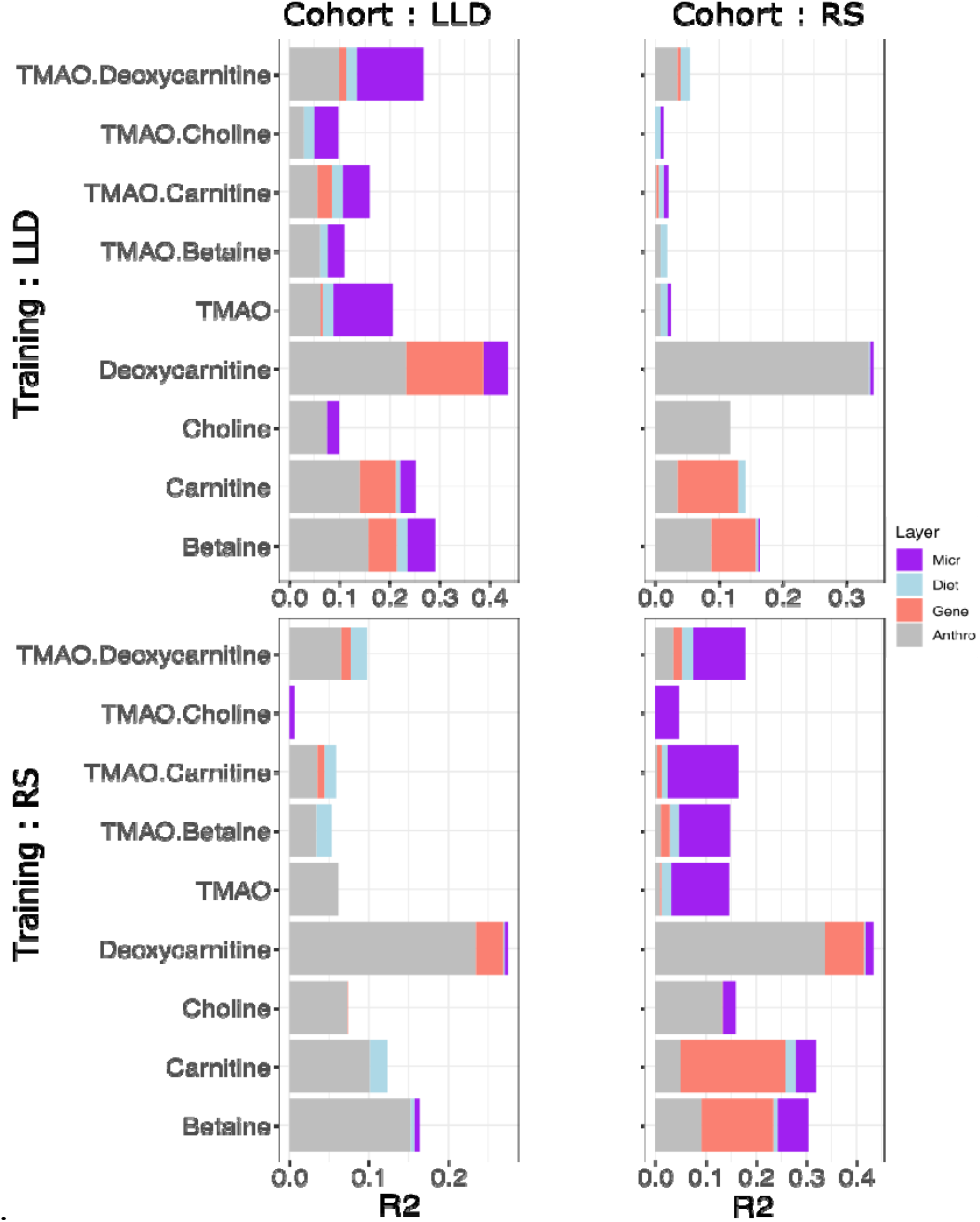
Variance explained in gut microbiome related metabolite levels and ratios by different data layers. X-axis shows the coefficient of determinations R2 gained with each additional data layer (gray: anthropometrics, red: genetics, blue: diet, purple: microbiome). Two models were trained, using LLD or Rotterdam Study III-2 (named RS) as the train set, and the left-out cohort as the test set respectively.

Subsequently, due to the lack of consistency found in the cohort-trained models, we sought to identify consistent associations between individual metabolites and host genetics, gut microbiome, or diet through a meta-analysis.

### Genetic variants are underlying levels of TMAO precursors, but not of TMAO itself

To evaluate host genetic determinants of TMAO-related metabolites and ratios, we performed genome-wide association study (GWAS) (N = 7,093). The results of the GWAS meta-analysis are illustrated in **Figure 3a**. The quantile-quantile plots indicated that any cryptic relatedness and/or population stratification were well-controlled after genomic correction (λ ranged between 1.00 and 1.02) (**Supplementary Figure 2, Supplementary Table 5**). Meta-analyses identified 55 independent genetic variants (see Methods section for description of independent genetic variant selection criteria) mapped to 5 genomic regions for betaine, 89 independent genetic variants mapped to 3 genomic regions for carnitine, 10 mapped to 3 genomic regions for choline and 37 mapped to 3 genomic regions for deoxycarnitine at Bonferroni corrected genome-wide significance level (*p*-value < 8.33×10^−9^) (**Supplementary Table 6, 7** and **8, Supplementary Figure 3**). Of these genomic regions, two genomics regions for betaine (6p21.1, 6q25.3), two for choline (2q34, 5q31.1) and two for deoxycarnitine concentration (10q21.2, 11p14.2) have not been reported in previous association studies (**Supplementary Table 7**). GWAS of TMAO revealed no genetic variant associated with TMAO concentration at genome-wide significant level (**Figure 3a**). Our findings are in line with previously published studies.^34, 35, 36, 37^ As this might suggest that genetic variants have a weak effect on variation in TMAO levels, we combined our results with the results from the publication of *Hartiala et al*^35^ in order to increase our sample size and improve power. However, no difference in signal was observed. Subsequently, we have also explored the genetic variants in the FMO region (**Supplementary Figure 4**). Even tough, previous studies reported a link between rare mutations in the *FMO3* gene and TMAO levels,^38^ no association (p-value > 0.1) was observed neither with common nor rare genetic variants of this region in our study. On the other hand, GWAS of TMAO to precursor ratio revealed genetic loci associated with TMAO to betaine ratio (n=2), TMAO to carnitine ratio (n=2) and TMAO to deoxycarnitine ratio (n=1) (*p*-value < 8.33×10^−9^) (**Figure 3b, Supplementary Table 9**). These genetic loci were identified in our GWA analysis of individual metabolites.

**Figure 3.**
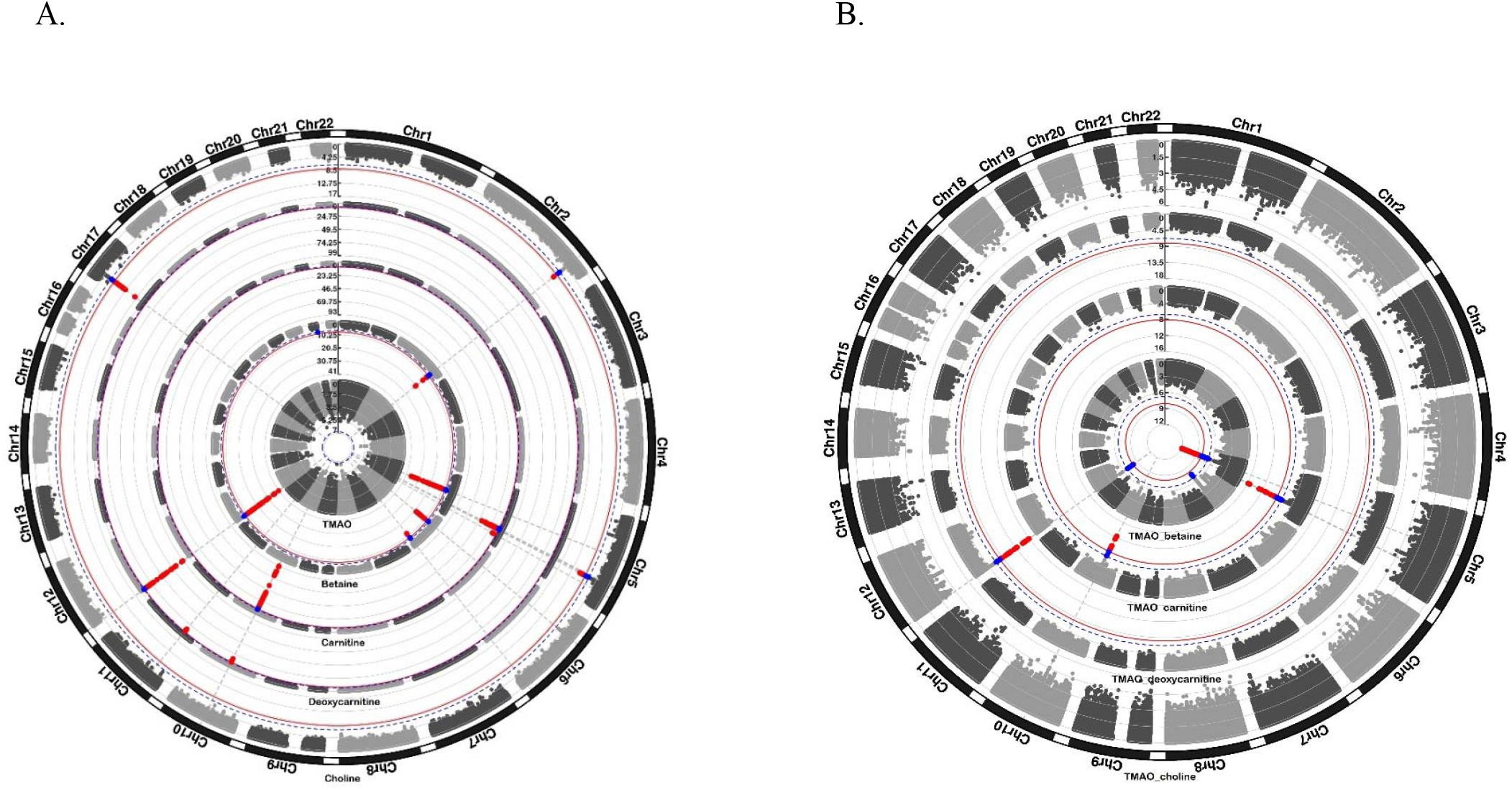
Results of genome-wide association analyses for individual metabolites (panel A) and TMAO-to-precursor ratio’s (panel B). Each dot represent a genetic variant. Genetic variants surpassing Bonferroni corrected significance threshold (*p-*value < 8.33×10^−9^) are highlighted in red. Genetic variants showing suggestive evidence of association (*p-*value < 1.7×10^−7^) are highlighted in blue.

The genetic variants found have previously been associated with various metabolic (e.g. plasma cholesterol and triglycerides levels, metabolite levels, liver enzyme levels), anthropometric (e.g., height, waist circumference, BMI) and medical traits (e.g., chronic kidney disease, diabetes mellitus, blood pressure). The list of associations for independent significant genetic variants (please see Methods for details) and all genetic variants in LD with these is provided in **Supplementary Table 10**.

Furthermore, we performed a sex-stratified analysis to capture sex-based differences (N_females_ = 4,026, N_males_ = 3,067). All significant associations observed in males and almost all significant associations found in females were also significant in the overall analysis (**Supplementary Figure 5, Supplementary Table 11**). An exception was the intronic variant mapped to *RP1* gene which showed a significant association with TMAO in females (beta = -0.29, *p-*value = 2.63×10^−9^) but not in males (beta = 0.02, *p-*value = 0.78) or in the overall analysis. This variant showed heterogeneity between males and females (*p-*value = 3.35×10^−5^). The *RP1* gene has been reported to function in photoreceptor differentiation (GeneCards Version 3: the human gene integrator). A gene-based association analysis revealed 11 genes associated with betaine, 16 with carnitine, 6 with deoxycarnitine and two genes with choline at gene-wide significance level (*p-*value < 4.42×10^−7^) (**Supplementary Table 12**). These genes were compared to previous association studies and an overview of both known genes replicated in our analysis and novel genes is shown in **Supplementary Table 12**. Several of the specific genes are considered in detail in the Discussion. No significant gene-sets were identified in the gene-set analysis (**Supplementary Table 13)**.

### Heritability estimates and genetic correlation

SNP-based heritability of metabolites was estimated in a range from 0.16 (SE = 0.07) for choline to 0.28 (SE = 0.08) for betaine using LD score regression (**Supplementary Figure 6**). An overlap of lead genetic loci was observed between betaine and choline (2q34), betaine and deoxycarnitine (12p13.33), carnitine and deoxycarnitine (10q21.2) and carnitine and choline (5q31.1) (**Supplementary Figure 7**). Additionally, we examined genetic overlap on a genome-wide level by computing genetic correlations. Evidence of suggestive genetic overlap was observed for TMAO and choline (ρ_genetic_□=□0.63, *p*-value□=□2.77□×□10^−2^) and betaine and choline (ρ_genetic_□=□0.54, *p*-value□=□5.7□×□10^−3^), while evidence of significant genome-wide genetic overlap was observed between TMAO and TMAO-to-precursor ratios (**Supplementary Table 14**).

### Microbial taxa are associated with plasma levels of TMAO, but not with those of its precursors

The results of the association analysis between the metabolites studied and 241 gut microbial taxa abundance are shown in **Figure 4**. There were 68 associations that surpassed the significance threshold after Bonferroni multiple test correction (*p*-value < 6.21×10^−5^). Significant associations were predominantly seen for TMAO (11/68) and TMAO-to-precursor ratios (56/68) (**Supplementary Table 15**). From those, the TMAO/carnitine ratio was associated with abundance of 23 microbial taxa and the TMAO/choline ratio with 15 microbial taxa. The top associated taxa corresponded to several *Ruminococcaceae* genera (NK4A21, UCG003, UCG005) which were positively associated to TMAO abundance ratio to its precursors, carnitine and choline. Other top associations included the class *Actinobacteria*, which was consistently negatively associated with TMAO-precursor ratios. The only significant association not directly related to TMAO was a positive association between the genus *Haemophilus* and betaine. Some members of this taxonomic group are able to oxidize choline to generate betaine, although we did not observe a negative association between these taxa and plasma choline.^39^ Association effect sizes across cohorts were generally concordant, with no evidence for heterogeneity in most associations (nominally significant heterogeneity in 120/2,169 associations and 4/64 among significant associations) (**Supplementary Table 15**). Interestingly, most of the significant or close to significant TMAO associations showed a different direction of effect than associations to at least one precursor (**Supplementary Figure 8**). This might highlight taxa with the potential to metabolize TMAO-precursors into TMA.

**Figure 4.**
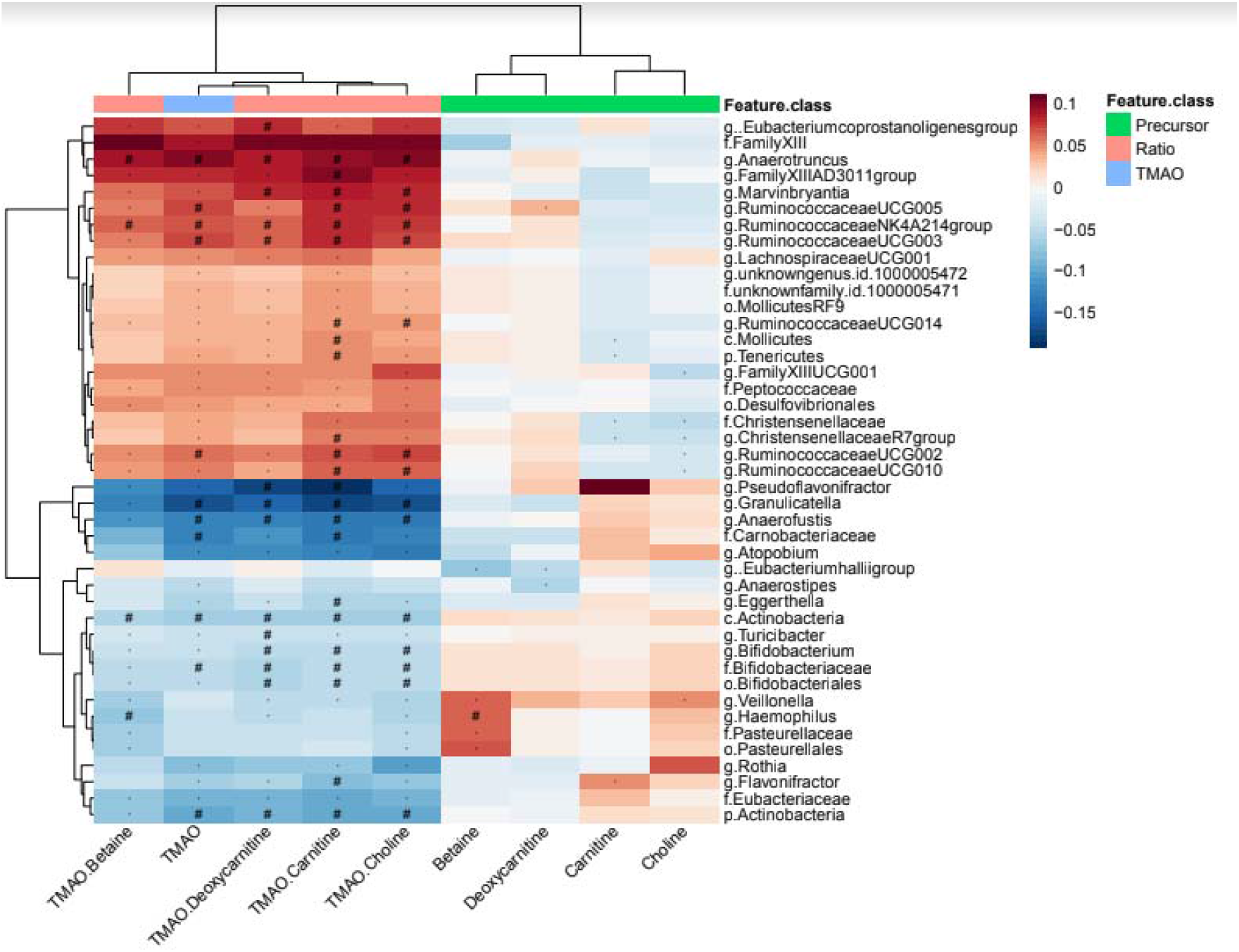
Heatmap showing results of the association analysis between metabolites and gut microbial taxa. Displayed results are after adjustment for age, sex, BMI, and study-specific covariates. Metabolites are displayed on x-axis and gut microbial taxa are shown on y-axis. Red color denotes positive associations and blue color stands for negative associations. Hash symbol (#) represents the Bonferroni significant associations (*p-*value < 6.2×10^−5^), while star denotes suggestive associations (*p-*value < 1.2×10^−3^).

Next, we performed such analyses using female (**Supplementary Table 16**) and male (**Supplementary Table 17**) stratified samples. In males we identified only four significant results, while we found 15 in females (**Supplementary Figure 9**). Most of these associations were for TMAO-precursor ratios, while only three of them were seen for TMAO (in females). If we focus on suggestive associations in one of the stratified analyses based on sex (*p*-value < 1.2×10^−3^) but not found to be associated in the overall analysis (*p*-value > 1.2×10^−3^), we could identify 14 associations, 10 of which showed a significant heterogeneity (*p*-value < 0.05) between females and males (**Supplementary Figure 10)**. The most noteworthy heterogeneous association is between *Coprococcus* and carnitine, for which a clear negative association is seen in males (beta = -0.0178, p-value = 0.039), which does not appear in females (beta = 0.006, *p*-value = 0.583). This taxon has previously been linked to be sex-biased in mammals, such as mice or pigs.^40, 41^

In order to examine causality between the microbial taxonomy and metabolite levels, we performed two-sample Mendelian Randomization (MR) between taxa and metabolites (excluding ratios). However, we failed to find any significant association (**Supplementary Table 18**).

### Diet is associated with plasma levels of TMAO and its precursors

Correlations between metabolites and dietary data categorized into 13 major food groups while adjusting for age, sex and BMI are presented in **Supplementary Figure 11** and **Supplementary Table 19**. There were 16 correlations that surpassed Bonferroni corrected threshold for multiple testing (*p-*value < 7.58 × 10^−4^). Top positive correlations were observed between TMAO or TMAO-to-precursor ratios and fish intake. Levels of circulating precursors, showed association with other food groups. Betaine levels were positively associated with grains, vegetables and nuts, while carnitine levels showed positive association with meat and negative correlations were observed between cheese intake and choline and eggs intake and deoxycarnitine. Overall, our findings are in line with literature findings.^11, 30, 42, 43, 44^

## DISCUSSION

We have performed an in-depth study to identify potential relationship between plasma levels of TMAO and its precursors betaine, carnitine, deoxycarnitine and choline on one hand and clinical outcomes, host genetics, gut microbiota composition and diet on the other hand in up to 7,834 participants from five prospective cohort studies mainly based on the general Dutch population. We found a statistically significant association between choline and higher risk of CVD. Furthermore, we have found that variation in gut microbiota and dietary composition were important determinants of TMAO levels and that host genetics and dietary composition were drivers of precursor levels. Among these, we identified several novel gut microbial drivers of TMAO levels and novel genetics determinants of TMAO precursor levels.

We observed a significant association of the circulating choline and higher CVD risk. Higher circulating levels of choline correlated with a 17% increase in risk for CVD events. Furthermore, circulating levels of choline showed a trend towards nominal association with all-cause mortality. Choline is an essential nutrient that plays a role in various metabolic process such as C1 metabolism and synthesis of phospholipids. As such, choline metabolism interacts with the pathways of insulin sensitivity, fat deposition and energy metabolism.^45^ The relationship between dietary choline intake and circulating levels of free choline and its metabolites is concealed by homeostatic regulations and rapid tissue uptake.^46^ As a result, the concentration range of choline and its metabolites is often quite narrow.^46^ Despite being a common dietary compound, we did not identify a strong predictive potential of diet in plasma choline levels. This might be related to the fact that blood samples in our study were mainly collected after overnight fasting. Based on previous literature findings significant increase of choline concentration was observed up to the 8h after intake.^46^ Previous studies reported associations of circulating levels of choline with higher risk of CVD, mortality and with some traditional cardiovascular risk factors including lower HDL, higher systolic pressure, triglycerides.^4, 8, 17, 47, 48^ However, there are also studies focusing on dietary choline intake with conflicting results. Higher dietary intake of choline was not predictive for incident coronary heart disease or CVD mortality.^28, 29, 30, 31^ A systematic review and meta-analysis of six prospective studies also reported no association of dietary choline intake and incident CVD.^49^ However, there was significant heterogeneity among those studies. Therefore, inconsistency in findings may be related to differences in dietary patterns, sample sizes, follow-up periods and geographical location. We also investigated the causality of our association by means of a MR analysis, which failed to support causal relationship. These findings are in line with previous literature findings.^50^ Future studies should focus on improving strength of the instrumental variables for plasma levels of choline.

In contrast to circulating choline, we did not find association between plasma TMAO levels and CVD risk or mortality in our population-based cohorts. Even though a number of studies have demonstrated a relationship between TMAO levels and CVD risk, these results are derived from individuals with high risk of CVD, existing disease or multimorbidity.^6, 7, 8, 51, 52, 53, 54^ In line with our results, evidence from other studies based on a general population is mainly suggesting that there is no association between TMAO and cardiometabolic markers, carotid intima media thickness, CVD events including heart attack and stroke and mortality.^9, 21, 55^ Taken together with our results, these findings suggest that circulating TMAO may not be a metabolite of concern in individuals from the general population. ^9, 17, 21, 55^

Next, we aimed to understand what factors are the most important drivers of metabolite variability in the general population. We estimated the amount of variability in circulating TMAO and its precursors explained by genetics, gut microbiome and diet. It is important to highlight that only linear additive effects were included and thus our estimates might be underestimation of the total variability explained. However, this analysis allowed us to compare the additive predictive power among the different data sources. Although the total variability of plasma TMAO levels that we could address is relatively small, a high proportion of that variability might be attributed to gut microbiome taxonomy. However, the results indicate a cohort-specific microbial contribution to TMAO levels. Similarly, the genetic effect on metabolite composition seems to differ between LLD and Rotterdam Study III-2. These heterogeneous results might be related to different demographics between the cohorts. For instance, the age breadth of LLD participants is wider (18-83 years) than Rotterdam Study III-2 (52 to 93 years), which is highlighted by the amount of variability explained by anthropometric factors seen in LLD in comparison to Rotterdam Study III-2. This demographic difference might be quite relevant in the metagenomic analysis, since our 16S data only allowed for resolution at the genus level, and important species and subspecies differences might be related with age.^56^ This heterogeneity drove us to perform meta-analysis of the different information layers available to identify common metabolite drivers among the different cohorts included.

Exploring the role of host genetics in underlying plasma levels of TMAO and its precursors revealed genome-wide significant variant associations with plasma levels of TMAO precursors but not for TMAO itself. We confirmed some of the previously reported associations implicated in determining plasma levels of all TMAO precursors. The *DMGDH, BHMT*, and *BHMT2* genes mapped to the 5q14.1 region have previously been linked to betaine levels (**Supplementary Table 9 and Figure 4**).^57, 58^ These genes are involved in betaine metabolism which is related to a series of interlinking metabolic pathways that include the methionine and folate cycles.^59^ Choline is also related to this pathway and we confirmed association with the *PEMT* gene (17p11.2) encoding an enzyme critical in phosphatidylcholine synthesis.^60^ Furthermore, the *SLC6A13* gene mapped to12p13.33 locus has previously been linked to deoxycarnitine levels while *SLC16A9* gene mapped to 10q21.2 locus has been associated with carnitine levels.^61^ Interestingly, we have identified 10q21.2 region as a novel genetic region underlying deoxycarnitine levels. The lead variant of this region mapped to the *SLC16A9* gene which is involved in urate metabolism. *SLC16A9* encodes a membrane transporter and is expressed in the intestine (GTEx Analysis Release V8), which might indicate a role in deoxycarnitine absorption. Previous studies showed that deoxycarnitine is an intermediary metabolite produced from carnitine by gut microbiota.^62^ However, deoxycarnitine may also act as an intermediate precursors to endogenous carnitine synthesis.^62^ Interestingly, we identified a novel genetic locus in the GWAS of deoxycarnitine that depicts this process. More specifically, the top lead intronic variant of 11p14.2 locus was mapped to *BBOX1* gene which it known to catalyzes the formation of carnitine from deoxycarnitine and is therefore involved in the carnitine synthesis pathway.

Additionally, we identified the 6p21.1 region as a novel region associated with betaine levels. This region has previously been associated with stroke and type 2 diabetes.^63, 64^ Genetic variants in linkage disequilibrium (r^2^ > 0.8) with our lead variant were associated with differential expression of *GNMT* and *PEX6* genes. Interestingly, *GNMT* gene is involved in a metabolism of methionine. Among novel regions, we have also identified 2q34 locus underlying the plasma levels of choline. Lead genetic variant of this region is mapped to *CPS1* gene which is involved in the urea cycle. Variants in this gene have previously been linked to creatinine, glycine, betaine and homocysteine levels, BMI, systolic blood pressure and cholesterol levels.^58, 65, 66, 67^

Although we were not able to detect genetic variants underlying plasma TMAO levels, we estimated that 20% of genetic variability in TMAO levels could be explained by common genetic variants. To discover genetic determinants of TMAO, future studies should further increase sample size and focus on complex genetic effects. Additionally, diet intervention studies might be of help, as these could decrease the variability attributed to gut microbiome and diet.

Microbial abundance was mainly associated with TMAO and TMAO to precursor ratios, which may be interpreted as a proxy for microbial conversion rates. Dietary precursors, on the other hand, did not show strong microbial associations. Several of the taxa we identified have previously been linked to TMAO. For instance, among the taxa belonging to the positively associated cluster, *Ruminococcus* or uncultured *Ruminococcaceae* have frequently been described to correlate with TMA and/or TMAO levels in mice and human.^17, 18, 20, 68, 69^ A member of the *Family III* was associated with TMAO levels in mice.^70^ *Anaerotruncus* was seen to be decreased upon resveratrol treatment in a mouse model and has been linked to TMAO metabolism.^68^ In the same study, several bacterial taxa were increased after resveratrol treatment, including *Bifidobacterium, Bifidobacteriaceae* and *Bifidobacteriales*, which are negatively associated with TMAO in the present study, also in agreement with other observations in humans.^12^ Interestingly, the strongest negative associations were found between TMAO to precursor ratios and *Pseudoflavonifractor* or *Granulicatella* genera, have not been reported before. However, both taxa have been linked to CVD, although in opposite directions.^71, 72^ *Pseduoflavonifractor* may produce a cholesterol derivative, coprostanol, which might be advantageous in the context of CVD, since this compound is not easily absorbed from the gut.^71^ On the other hand, *Granulicatella* was found among the bacterial changes linked with a severe coronary artery disease.^72^ Conversely, other taxa that are often linked to TMAO^73^ did not show any significant association in our study, including *Clostridia* or *Escherichia* genera. *Desulfovibrionales* although did not pass the Bonferroni corrected *p-*value threshold showed consistent positive associations with TMAO.

Although we found compelling associations between TMAO and bacterial taxa that were for the most part opposite or absent for its precursors, in-silico causal inference of TMAO metabolism from the associated taxa by means of two-sample MR was not successful. The weak microbial instruments may have added noise to this analysis, preventing the identification of consistent signals. Also, the lack of strong TMAO genetic architecture complicates the analysis. Thus, further experimental analyses will be needed to probe a causal relationship between the bacteria reported here and TMAO metabolism.

In addition to gut microbiota, diet composition was observed to be an important determinant of metabolite levels. For instance, TMAO levels and TMAO to precursor ratios showed positive association with fish intake. Previous studies linked TMAO levels to fish intake and this association has been demonstrated to vary throughout populations and/or regions.^11^ Fish consumption was associated with TMAO levels in Asian countries and some European countries, while intake of eggs and red meat showed stronger correlation with TMAO in the population of United States.^14, 15, 16, 74^ In our study population, no association was found between TMAO and eggs or red meat.^21^ Fish and seafood are rich in TMAO and TMA and they can be directly absorbed without being transformed by gut microbiota.^14^ As previous studies linked TMAO to atherosclerosis and CVD, these findings might be contra intuitive as fish is generally accepted to be cardioprotective.^75^ The risk of CVD is not increased with fish consumption as fish contains cardioprotective molecules such as ω3-poly unsaturated fatty acids.^76^ Previously published randomized trials also indicated that diet with a large proportion of lean white fish (rich in TMAO) reduces risk of CVD.^75^

Overall, the large sample size, population-based design and comprehensive molecular and epidemiological data of our study helped us to investigate the sources of variation of TMAO and its precursor metabolites in the general population and their health-related consequences. We were able to study not only TMAO but also the compounds implicated in the TMAO biosynthesis pathway. Furthermore, we were also able to improve statistical power and internally cross-check the findings by combining data from five population-based studies. However, our study also has limitations. The gut microbial composition was determined from fecal samples that might not be representative of the overall gut microbial composition.^77^ In addition, within-species genetic variation is known to modulate bacterial-related metabolites, thus the taxonomic resolution of 16S might not properly reflect the metabolic potential of the present strains.^23^ Metagenomic-shotgun sequencing experiments will be needed to address that level of variation. The cross-sectional nature of our metabolomics measurements and gut microbiota assessment in our study only allowed us to investigate the relationship between the two at one time point. To complement our findings and advance our understanding, future studies should focus on assessing longitudinal changes. For instance, a 1-year follow-up study in the general population reported large variation in plasma TMAO concentrations, which might underlie the heterogeneous associations related to this metabolite.^78^ Since our study population comprised of individuals of European origin and as some of associations might be population specific, expanding research in non-European populations will be needed.

In conclusion, our data adds up to the mounting evidence of research showing that TMAO is not associated with an increased risk of CVD in general population, despite earlier evidence suggesting this to be the case among patient groups. However, we did show a significant relation between plasma choline levels and higher CVD risk. Our MR revealed no evidence for a causal link between TMAO or its precursors with incident CVD. Furthermore, we also identified several determinants explaining the variability of TMAO and its precursors blood levels in humans. Gut microbiome was mainly associated with TMAO-to-precursor ratios, although the total variability explained of TMAO concentration remains mild and cohort specific. Diet was associated with both TMAO and its precursors but could not explain a great proportion of their variation. Genetic contributions to precursor concentrations were greater than to TMAO itself, where no strong genetic effects were seen. The biological mechanisms underlying these associations should be the subject of further studies.

## METHODS

### Study population

Our study population included 7,834 participants from five cohort studies. Detailed description of participating studies can be found in **Supplementary Material** and descriptive characteristics of study participants are shown in **Table 1**. Each study was approved by ethical committees (please see **Supplementary Material** for details). Written informed consent was obtained from all participants.

### Metabolite profiling

TMAO and its precursors betaine, carnitine, deoxycarnitine and choline were quantified in plasma samples of participates from five cohorts by using liquid chromatography tandem mass spectrometry (LC-MS/MS) method. Detailed description of the method can be found elsewhere.^33^ Briefly, before introducing the sample to the mass spectrometer, an analytic column with a C18 stationary phase as used to realize an on-line cleanup of it. The analytes were not retained by this stationary phase but important matrix interferences were retained, such as (phosphor-)lipids.^33^ The descriptive statistics of metabolites were coherent across the cohorts (**Supplementary Table 1**). In addition to individual metabolites, ratios of TMAO to its dietary precursors were also calculated.

### Incident clinical outcomes

The Rotterdam Study I-4, Rotterdam Study III-2 and LLS cohorts had data on incident clinical outcomes including cardiovascular disease (CVD) and mortality. Incident CVD events were defined as incident stroke, myocardial infarction, angina pectoris and heart failure according to the codes of International Classification of Disease, 10^th^ edition. Incident CVD events were assessed continuously through an automated digital linkage of study database to medical records maintained by general practitioners in the Rotterdam Study and from general practitioner records in the LLS.^79, 80^ Information on vital status is additionally obtained from the central registry of the municipality of the city of Rotterdam and in January 2021 vital status for participants of Leiden Longevity Study was updated through the Personal Records Database (PRD) which is managed by Dutch governmental service for identity information (https://www.government.nl/topics/personal-data/personal-records-database-brp).^81^

### Baseline clinical characteristics

The baseline clinical characteristics were obtained by means of interview, physical examination, blood sampling or medical records from general practitioner. Assessment of current smoking status, weight, height, blood pressure, glucose levels, total cholesterol in low-density lipoproteins (LDLC) and high-density lipoproteins (HDLC), serum triglycerides (TG), creatinine, and medication use including lipid-lowering medication and use of medication indicated for the treatment of diabetes. Diabetes was defined as fasting glucose levels above 7Lmmol/L, non-fasting glucose levels above 11.1 mmol/L (only if non-fasting levels were unavailable) or use of medication indicated for the treatment of diabetes,^82^ or through medical records from general practitioner. Hypertension was defined as a systolic blood pressure ≥140□mmHg, diastolic blood pressure ≥90□mmHg, or use of medication for the treatment of hypertension,^82^ or through medical records from general practitioner. BMI was calculated as weight in kilograms divided by square of heights in meters. Quantified creatinine was used to calculate the estimated glomerular filtration rate (eGFR) using the Chronic Kidney Disease Epidemiology Collaboration equation (CKD-EPI).^83^

### Genotyping and imputation

Details on genotyping platforms, calling method and quality control procedures in participating studies are shown in **Supplementary Table 20**. Commercially available genotyping arrays were used for genotyping. Similar quality control procedures were applied in each study prior genotype imputation (**Supplementary Table 20)**. Genotypes in each cohort were imputed by Haplotype Reference Consortium (HRC) reference panel on a Michigan Imputation Server.^84^

### Microbiome processing

The Rotterdam Study III-2 and LLD cohorts had available 16S-amplicon sequence data from fecal samples matching plasma collection. Fecal sample collection and 16S sequencing was described elsewhere.^85, 86, 87^ In brief, DNA was isolated from the fecal samples belonging to the Rotterdam Study III-2 cohort and the V3 and V4 variable regions of the 16S rRNA gene were amplified and sequenced on Illumina MiSeq sequencer. Similarly, DNA was isolated from fecal samples of LLD participants and the 16S V4 region was sequenced at the Broad Institute using Illumina MiSeq. 16S rRNA data was processed as previously described in a large 16S meta-analysis including both cohorts.^88^ In brief, samples were rarified to 10,000 reads. Reads were classified to a given taxonomic level (genus to phyla) using RDP classifier (v2.12).^89^ Reads below 0.8 posterior probability to belong to a given taxonomic level were discarded. For each sample, each taxonomic level was centered-log ratio transformed (clr). Only taxa observed in above 10% of participants per cohort were used for association resulting on 241 microbial taxa.

### Dietary assessment

The Rotterdam Study cohorts, LLS and LLD had data on dietary intake collected via validated food frequency (FFQ) questionnaire. Data on dietary intake in the LLS, LLD and Rotterdam Study cohorts were collected via validated FFQs.^90, 91, 92^ The FFQs assess the frequency of consumption of food items and the number of servings per day. Additionally, information on portion size, type of food item, and preparation methods was collected. The average daily energy and nutrient intake was calculated using the Dutch Food Composition Database. Specific food items were aggregated into food groups in grams per day. The major food groups overlapping between the cohorts were used for subsequent analysis including vegetables, fruit, grains, nuts, eggs, fish, meat, poultry, processed meat, cheese, milk, yoghurt and total dairy products.

## Statistical analyses

### Incident clinical outcomes analysis

Metabolites were transformed using rank-based inverse normal transformation. The relationship between metabolites and incident clinical outcomes was assessed using Cox proportional hazards regression models with age as time scale. The analyses were adjusted for sex, BMI, hypertension, diabetes, LDLC, HDLC, TG, lipid-lowering medication use, current smoking and fasting status (if appropriate) (model 1). Subsequently, the associations were adjusted for eGFR (model 2). The proportional hazard assumption was checked using statistical tests incorporated in the *survival* package. Violation of this assumption was observed for some of the covariates and was resolved by stratification. In the LLS, Cox-type random effect (frailty) regression models was used in order to adjust for family relations. All analyses were performed using R. The summary statistics results of participating studies were combined using inverse variance-weighted fixed-effect meta-analysis in METAL.^93^ The heterogeneity of effects was assessed by I^2^ which indicates the percentage of variance in the meta-analysis attributable to study heterogeneity.^94^ To model correlation between metabolites, we first used the method of Li and Ji to calculate number of independent tests.^95^ The Bonferroni corrected significance threshold was calculated based on the number of independent tests and set at 0.05/6 independent metabolites = 8.33×10^−3^. Additionally, all analyses were stratified by sex. The same steps were followed as for overall analysis.

### Incident clinical outcomes-metabolites Mendelian Randomization analysis

The potential casual effect of metabolites on clinical CVD outcomes was assessed by MR analysis using TwoSampleMR package. GWAS summary statistics results were obtained from a large meta-analysis comprising coronary artery disease cases and controls of UK Biobank resource and CARDIoGRAMplusC4D.^96^ Genetic variants with *p-*value < 1×10^−5^ were used as instruments. Independent genetic variants were selected based on r^2^ in European reference data. The results were kept if these were based on at least three shared genetic variants. Causality was estimated using various MR method’s including Inverse variance weighted (IVW), MR-Egger, Wald ratio, Weighted median, Simple Mode and Weighted Mode.

### Estimation of determination coefficient in different data layers

The Rotterdam Study cohorts and LLD were used to estimate the total determination coefficient (R^2^) from each of the analyzed data layers in each of the metabolites or metabolite ratios. We used features present in both cohorts including anthropometric covariates (age, sex, BMI), 8 overlapping dietary items, 242 bacterial taxonomic abundances and the number of suggestive genetic variants (*p-*value < 1×10^−5^) from a meta-analysis of the GWAS results from LLS and 300-OB. These two cohorts were used for preselecting variants and were not used to train the model or estimate R^2^. Taxa-abundance was clr-transformed, while metabolites and diet were inverse-rank normal transformed.

We trained a regularized additive linear model, elastic net (glmnet v4.0), and selected the best combinations of hyperparameters alpha (regularization mix) and lambda (regularization strength) through a 5-repeated 10-fold cross-validation procedure using the root mean square error (RMSE) as performance metric (caret v6.0, tunelength=10). We trained a model for each metabolite (or metabolite ratio) using two different training sets, a training set consisting of the LLD cohort (784 samples) and a training set consisting of the Rotterdam Study III-2 cohort (772 samples). For the test set (LLD in the Rotterdam Study III-2-trained or the Rotterdam Study III-2 in LLD-trained model), the determination coefficient (R^2^) was estimated in nested models. To estimate anthropological R^2^, all other coefficients were made 0. To estimate genetics R^2^, all non-genetics, non-anthropological covariate coefficients were made 0. This was followed by the addition of non-0 diet parameters and finally the complete model including microbial features. Individual layer R^2^ was quantified by subtracting the R^2^ from the nested models, e.g microbial R^2^ was estimated by subtracting the complete model R^2^ and the diet model.

### Genome-wide association analysis

Each participating study performed genome-wide association analysis under an additive model using metabolites as dependent variable and variant allele dosage as a predictor. Prior to the analysis, metabolites were transformed using rank-based inverse normal transformation. The association analysis was adjusted for age, sex, fasting status if applicable, familial relatedness if appropriate and principal components if needed. Study-specific details on covariates and software used to run the analysis are provided in **Supplementary Table 20**. The quality control (QC) was performed using a standardized protocol provided by Winkler et al. ^97^ Genetic variants with minor allele count below 10 and low imputation quality (r^2^ < 0.3) were excluded. The summary statistic results were combined using fixed-effect meta-analysis in METAL.^93^ To account for a small amount of population stratification or unaccounted relatedness, genomic control was applied. After meta-analysis, genetic variants that were present in less than three participating studies were filtered out. The Bonferroni corrected genome-wide significance threshold was set at 5×10^−8^/6 independent metabolites = 8.33×10^−9^. Additionally, the analyses were stratified by sex. The same QC steps were followed as for overall analysis. The sex-stratified summary statistic results were combined using fixed-effect meta-analysis in METAL while applying genomic control.^93^ Test statistics of each variant were tested for heterogeneity between males and females.

### Functional mapping and annotation

Functional Mapping and Annotation of genetic associations (FUMA) was used to characterize genomic loci.^98^ Genetic variants that passed Bonferroni corrected genome-wide significance threshold and were independent from each other (r^2^ < 0.6) were defined as independent genetic variants. Independent significant genetic variants with r^2^ < 0.1 were defined as lead genetic variants. Independent significant genetic variants with r^2^ ≥ 0.1 or that were 250 bp or closer were assigned to the same genomic risk locus. Each locus was represented by the top lead genetic variant with minimal *p*-value in the locus. Functional annotation was performed using Combined Annotation Dependent Depletion (CADD)^99^, HaploReg^100^, and RegulomeDB^101^ tools as implemented in FUMA.

### Gene based and gene set enrichment analysis

Genome-wide summary statistics were used to perform gene-based analysis using MAGMA as implemented in FUMA.^98^ Genetic variants were assigned to the genes from Ensembl build 85 based on genomic location. All genetic variants mapped to the protein coding genes were tested for association with metabolites using SNP-wide mean model. 1000G phase 3 was used as a reference panel to calculate LD across SNPs and genes. To account for multiple testing, Bonferroni correction was calculated and gene-wide significance threshold was set at 0.05/(18 861 tested genes × 6 independent metabolites) = 4.42×10^−7^. Subsequently, gene sets enrichment analysis was also performed using FUMA. Hypergeometric tests were performed to test if genes of interest are overrepresented in any of 15,496 pre-defined gene sets obtained from MsigDB. Multiple test correction was calculated based on the total number of gene sets (*p-*value = 0.05/15496 = 3.23×10^−6^).

### Heritability estimates and genetic correlation

Heritability of metabolites and genetic correlation between them were estimated from GWAS results using LD Score Regression approach.^102^ We used pre-computed LD scores for Europeans. Only genetic variants available in HapMap3 were used.

### Gut microbial taxa and metabolites association analysis

Each of the metabolites and TMAO-to-precursor ratios were rank-based inverse normal transformed to ensure normality. Standard linear regression models were carried out to associate the transformed metabolite and taxonomy abundances while adjusting for age, sex, BMI, and study-specific covariates (sample batch, time in mail and storage time in the Rotterdam Study III-2). This analysis was also reproduced in sex-stratified samples. Common taxonomy associations in the Rotterdam Study III-2 and LLD were meta-analyzed using a fixed-effects, inverse-variance analysis (R package meta v4.12). Association’s heterogeneity was measured by Cochran’s Q statistic.

In order to correct for multiple testing, we determined the number of independent tests using the method of Li and Ji.^95^ There were 134 independent tests among microbial taxa and 6 independent tests among metabolomics measures. The Bonferroni significance threshold was set at 0.05/(134 independent microbial taxa × 6 independent metabolites) = 6.2×10^−5^, while a suggestive threshold was set at 1/ (134×6) = 1.2×10^−3^.

### Gut microbial taxa and metabolites Mendelian Randomization analysis

Potential causal relation between gut microbial taxa and metabolite levels was tested by two-sample MR using the TwoSampleMR package. Only taxa to metabolite associations that surpassed our significant thresholds (*p*-value < 6.2×10^−5^) were considered for the analysis. We used the GWAS summary statistics for metabolites produced in this work, while for microbial taxa we obtained summary statistics from a large 16S meta-analysis comprising 18,340 individuals from 24 cohorts including both Rotterdam Study III-2 and LLD.^88^ Genetic variants with *p-*value < 1×10^−5^ were selected as instruments. Bacterial taxa with no available instruments were removed from the analysis. Independent genetic variants were selected as instrumental variables based on r^2^ threshold of 0.001 (1000 Genomes in the European reference population). The number of instruments varied between 11 and 20. The causality was estimated using various MR method’s including Inverse variance weighted (IVW), MR-Egger, Wald ratio, Weighted median, Simple Mode and Weighted Mode. In addition, we also assessed genetic variant heterogeneity and evidence of horizontal pleiotropy (using egger). Individual summary statistic for genetic variants were estimated using Wald ratio tests.

### Diet and metabolites correlation analysis

Each food group, metabolite and TMAO-to-precursor ratio were rank-based inverse normal transformed prior to analysis. Partial correlation coefficients were calculated between each transformed food group item and metabolite or metabolite ratio while adjusting for age, sex and BMI. Summary statistics results of participating studies were combined by performing fixed-effect meta-analysis in METAL.^93^ To model correlation between food groups, method of Li and Ji was used to calculate number of independent tests. Associations were considered significant if they surpassed Bonferroni corrected significance threshold of 0.05/ (11 independent food groups × 6 independent metabolite) = 7.58 × 10^−4^.

## Data Availability

The data is available from the author on request.

## Data availability

The data is available from the author on request.

## Acknowledgement

†We dedicate this paper to the memory of Prof. Marten Hofker. We thank all the participants in the Rotterdam Study, Leiden Longevity Study, Lifelines-DEEP cohort, and 300-OB cohort for their participation and the project staff for their help and management.

## Rotterdam Study

The Rotterdam Study is supported by the Erasmus MC University Medical Center and Erasmus University Rotterdam; The Netherlands Organisation for Scientific Research (NWO); The Netherlands Organisation for Health Research and Development (ZonMw); the Research Institute for Diseases in the Elderly (RIDE); The Netherlands Genomics Initiative (NGI); the Ministry of Education, Culture and Science; the Ministry of Health, Welfare and Sports; the European Commission (DG XII); and the Municipality of Rotterdam. The contribution of inhabitants, general practitioners and pharmacists of the Ommoord district to the Rotterdam Study is gratefully acknowledged. The contributions to the Management Team specifically and the Rotterdam Study at large of the following persons are pivotal to the daily operations of the study and highly appreciated: Jacobus Lubbe, Gabriëlle Bakker, Eric Neeleman, Jeannette Vergeer, Anneke Korving, Pauline van Wijngaarden, Jolande Verkroost – van Heemst, Silvan Licher, Isabel van der Velpen.

## LLD

This project is supported by the Netherlands Heart Foundation (IN-CONTROL CVON grant 2012-03 to T.H., M.G.N., L.A.B.J., F.K., A.Z., P.E.S., C.M.D. and J.F. and IN-CONTROL CVON grant 2018-27 to M.G.N., N.P.R., L.A.B.J., F.K., A.Z., and J.F.). F.K. is also supported by the Noaber Foundation. A.Z. is also supported by the European Research Council (ERC) Starting Grant 715772, Netherlands Organization for Scientific Research (NWO) VIDI grant 016.178.056, and the NWO Gravitation grant Exposome-NL (024.004.017). J.F. is also supported by an ERC Consolidator grant (grant agreement No. 101001678), a NWO-VICI grant VI.C.202.022, and the Netherlands Organ-on-Chip Initiative, an NWO Gravitation project (024.003.001) funded by the Ministry of Education, Culture and Science of the government of The Netherlands. M.G.N. is further supported by a European Research Council (ERC) Consolidator grant (3310372), an ERC Advanced Grant (FP/2007-2013/ERC grant 2012-322698), and a Spinoza Prize (NWO SPI 92-266). L.A.B.J. is supported by a Competitiveness Operational Programme grant of the Romanian Ministry of European Funds (HINT, P_37_762, MySMIS 103587).

## LLS

The LLS has received funding from the European Union’s Seventh Framework Programme (FP7/2007-2011) under grant agreement no. 259679. This study was supported by a grant from the Innovation-Oriented Research Program on Genomics (SenterNovem IGE05007), the Centre for Medical Systems Biology, The Netherlands Consortium for Healthy Ageing (grants 05040202 and 050060810), all in the framework of The Netherlands Genomics Initiative, Netherlands Organization for Scientific Research (NWO), and BBMRINL, a research infrastructure financed by the Dutch government (NWO 184.021.007 and 184.033.111), by the Netherlands CardioVascular Research Initiative (CVON201-03), VOILA (ZonMW 457001001) and Medical Delta (scientific program METABODELTA: Metabolomics for clinical advances in the Medical Delta). D.V. is supported by the Netherlands Consortium of Dementia Cohorts (NCDC), which is funded by Deltaplan Dementie from ZonMW Memorabel (project number 73305095005) and Alzheimer Nederland. The contribution of all participants of the Leiden Longevity Study is gratefully acknowledged.

## 300-OB

This project is supported by the Netherlands Heart Foundation (IN-CONTROL CVON grant 2012-03 to T.H., M.G.N., L.A.B.J., F.K., A.Z., P.E.S., C.M.D. and J.F. and IN-CONTROL CVON grant 2018-27 to M.G.N., N.P.R., L.A.B.J., F.K., A.Z., and J.F.). M.G.N. is further supported by a European Research Council (ERC) Consolidator grant (3310372), an ERC Advanced Grant (FP/2007-2013/ERC grant 2012-322698), and a Spinoza Prize (NWO SPI 92-266). L.A.B.J. is supported by a Competitiveness Operational Programme grant of the Romanian Ministry of European Funds (HINT, P_37_762, MySMIS 103587). NPR is recipient of a grant of the ERA-CVD Joint Transnational Call 2018 supported by the Dutch Heart Foundation (JTC2018, project MEMORY; 2018T093)

The research is supported by Medical Delta, scientific program METABODELTA: Metabolomics for clinical advances in the Medical Delta. This research was part of the Netherlands X-omics Initiative and partially funded by NWO, project 184.034.019.

Data on coronary artery disease / myocardial infarction have been contributed by the CARDIoGRAMplusC4D and UK Biobank CardioMetabolic Consortium CHD working group who used the UK Biobank Resource (application number 9922). Data have been downloaded from www.CARDIOGRAMPLUSC4D.ORG.

## AUTHOR CONTRIBUTION

Conception and design of the study: SAS, PES, CvD, JF, DV. Collection of the data: AK, MB, MG, MvF, ICLvM, MS, AH, TH, MAI, MK, TV, RK, MN, JHWR, NpR, AZ, FK, PES, CvD, JF. Analysis: SAS, SA, DV. Interpretation of the data: SAS, SA, AK, MB, TV, PES, CvD, JF, DV. Drafting of the manuscript: SAS, DV. All authors read, revised and approved the final draft.

