## Supplementary Material for "TMAO and its precursors in relation to host genetics, gut microbial composition, diet, and clinical outcomes: Meta-analysis of 5 prospective population-based cohorts"

Andreu-Sánchez **S** et al.

**Content**

**Description of study population**…………………………………………………………………………..3

**Supplementary Figure 1.** Phenotypic correlation between metabolites included in the analysis………...5

**Supplementary Figure 2.** Quantile-quantile plots for genome-wide association meta-analyses…………6

**Supplementary Figure 3.** Regional plots of lead genetic variants associated with circulating metabolite levels………………………………………………………………………………………………………..8

**Supplementary Figure 4.** Regional plot for the region encompassing FMO genes based on the GWAS summary statistics for TMAO…………………...………………………………………………………...11

**Supplementary Figure 5.** Results of sex-stratified genome-wide association analysis for single metabolites………………………………………………………………………………………………...12

**Supplementary Figure 6.** SNP-based heritability estimated using LD-score regression………………..13

**Supplementary Figure 7.** Overlap of lead genomic loci identified in the GWAS of TMAO and its precursors……………………………………………………………………………………………….…14

**Supplementary Figure 8.** Top associations of microbial taxonomy with metabolites..……………..….15

**Supplementary Figure 9.** Results of association analysis between metabolites and gut microbial taxa in males and females……...………………………………………………………………………………….16

**Supplementary Figure 10.** Gender-heterogeneous associations of microbial taxonomy with metabolites………………………………………………………………………………………………...17

**Supplementary Figure 11.** Results of correlation analysis between metabolites and ratios and food groups……………………………………………………………………………………………………...18

**References**………………………………………………………………………………………………...19

**Description of study population**

**Leiden Longevity Study (LLS)**

The Leiden Longevity Study (LLS) is a family population-based study that consists of 421 long-lived families of European descent and was enrolled between 2002 and 2006. Families were included if at least two long-lived siblings were alive and fulfilled the age criterion of 89 years or older for males and 91 years or older for females, representing <0.5% of the Dutch population in 2001.^1, 2^ In total, 944 long-lived proband siblings (mean age=94 years, range=89–104), 1,671 offspring (mean age=61 years, range=39–81) and 744 spouses thereof (mean age=60 years, range=36–79) were included.^3^ In this study, we used offspring and spouses. In accordance with the Declaration of Helsinki, we obtained informed consent from all participants prior to their entering the study. Good clinical practice guidelines were maintained. The study protocol was approved by the ethical committee of the Leiden University Medical Center before the start of the study (P01.113).

**LifeLines-DEEP (LLD)**

The LifeLines-DEEP cohort is a sub-cohort of LifeLines study, a prospective population-based cohort study in the north of the Netherlands established. The LifeLines cohort was established in 2006 among participants aged from 6 months to 93 years in order to gain insights into the etiology of healthy aging.^4^ At the baseline, the participants filled in extensive questionnaires and visited one of the LifeLines Research Sites twice for physical examinations. After completion of inclusion in 2013, the cohort includes 165,000 participants. A follow-up questionnaire was sent to each participant every 18 months and follow-up measurements of the health parameters were performed every 5 years.^5^ A subset of approximately 1500 LifeLines participants aged 18–81 years was included in Lifelines-DEEP. These participants were examined more thoroughly, specifically with respect to molecular data. Additional biological materials and information on health status were collected for these participants.^5^ The LifeLines-DEEP study is approved by the Ethical Committee of the University Medical Center Groningen.^5^ All participants provided written informed consent.

**Rotterdam Study (RS)**

The Rotterdam Study is a prospective, population-based cohort study among individuals living in the well-defined Ommoord district in the city of Rotterdam in The Netherlands.^6^ The aim of the study is to discover the causes of diseases and thereby identify potential targets for preventive interventions of cardiovascular, endocrine, hepatic, neurological, ophthalmic, psychiatric, dermatological, otolaryngological, locomotor, and respiratory diseases in mid-life and late-life. The cohort was initially defined in 1990 among 7,983 persons, aged 55 years and older, who underwent a home interview and extensive physical examination at the baseline and during follow-up rounds every 3-4 years (RS-I). Cohort was extended in 2000/2001 (RS-II, 3,011 individuals aged 55 years and older) and 2006/2008 (RS-III, 3,932 subjects, aged 45 and older). In summer of 2016, the recruitment of another extension started that targeted participants aged 40 years and over. The establishment of this extension is expected yield around 3000 new participants. The Rotterdam Study has been approved by the Medical Ethics Committee of the Erasmus MC (registration number MEC 02.1015) and by the Dutch Ministry of Health, Welfare and Sport (Population Screening Act WBO, license number 1071272-159521-PG). The Rotterdam Study Personal Registration Data collection is filed with the Erasmus MC Data Protection Officer under registration number EMC1712001. The Rotterdam Study has been entered into the Netherlands National Trial Register (NTR; www.trialregister.nl) and into the WHO International Clinical Trials Registry Platform (ICTRP; www.who.int/ictrp/network/primary/en/) under shared catalogue number NTR6831. All participants provided written informed consent to participate in the study and to have their information obtained from treating physicians.

**300 Obesity (300-OB) cohort**

The 300-Obesity (300-OB) cohort study consists of 302 individuals between 55 and 80 years of age with a BMI > 27 kg/m2 at screening. All participants were included between the year 2014 and 2016. This cohort is part of the Human Functional Genomics Project (HFGP) at the Radboud university medical center, which contains a collection of cohorts with various backgrounds. Most of these participants previously took part in the Nijmegen Biomedical Study – Non-Invasive Measurements of Atherosclerosis 1 (NBSNIMA1), a population-based survey of inhabitants of the municipality of Nijmegen. All participants signed an informed consent form prior to sample collection. The Ethical Committee of Radboud University Nijmegen approved the study. Details of the study population and cardiovascular parameters have been reported previously.^7, 8^

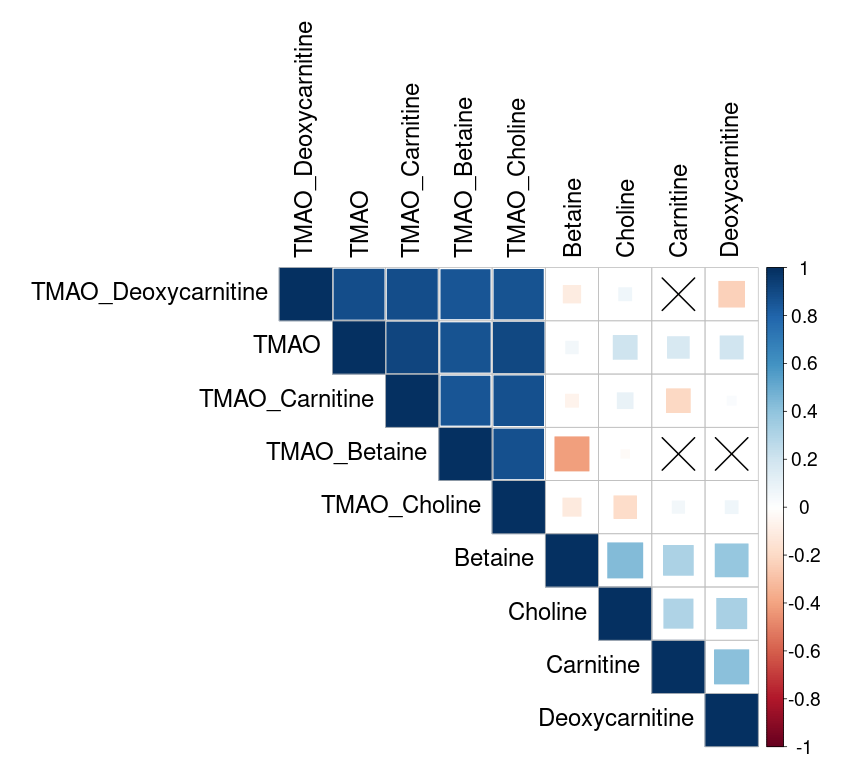

**Supplementary Figure 1.** Phenotypic correlation between metabolites included in the analyses. Displayed results are derived from the meta-analysis of correlation coefficients based on all five participating cohorts. Blue color stands for positive correlation and red color for negative correlation. The correlations with *p*-value > 0.05 are denoted with cross.

B. TMAO

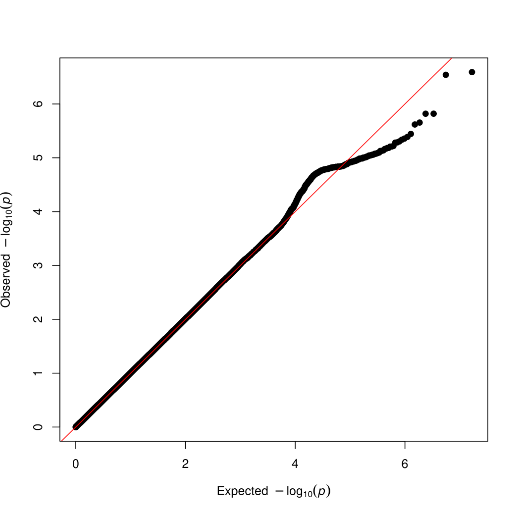

C. Betaine
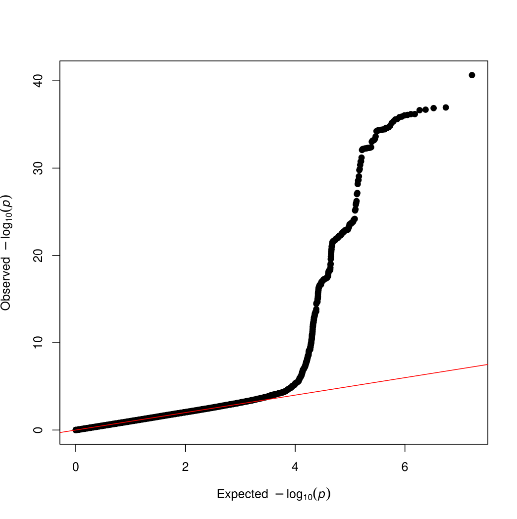

D. Carnitine
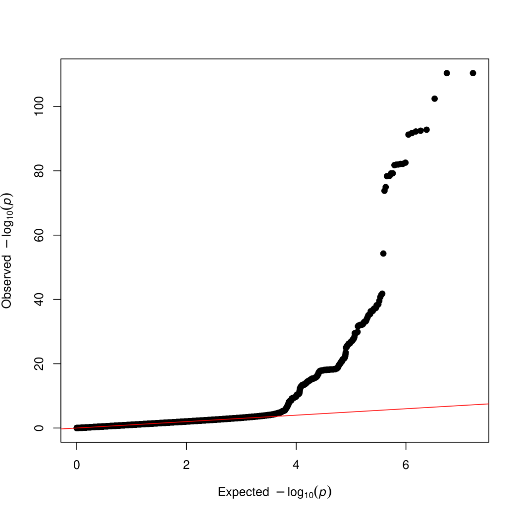

E. Deoxycarnitine

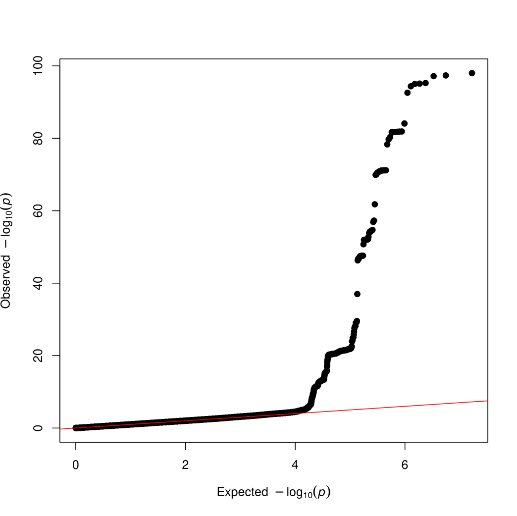

**Supplementary Figure 2.** Quantile-quantile plots for genome-wide association meta-analyses (1 of 2).

G. Choline

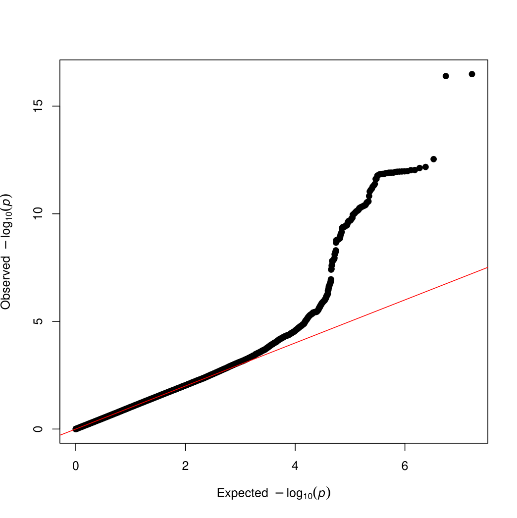

**Supplementary Figure 2**. Quantile-quantile plots for genome-wide association meta-analyses. (2 of 2).

B. **Betaine (5)**

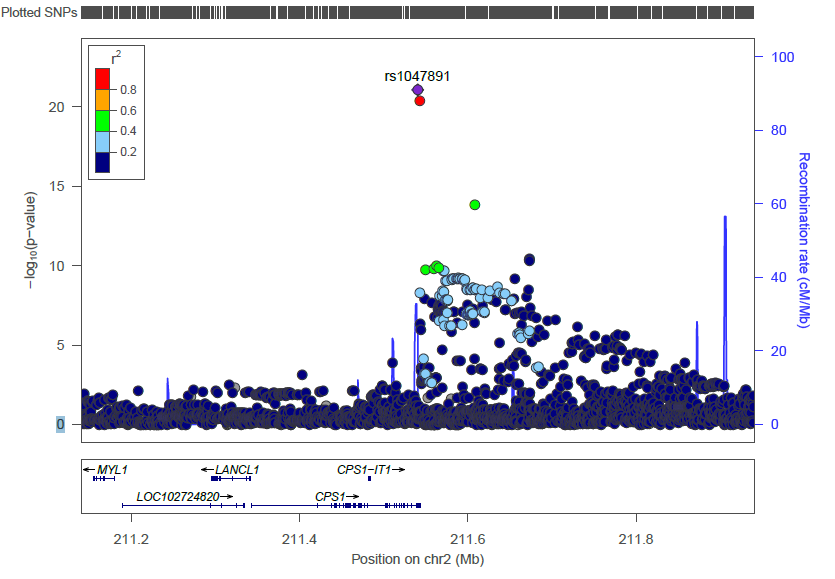

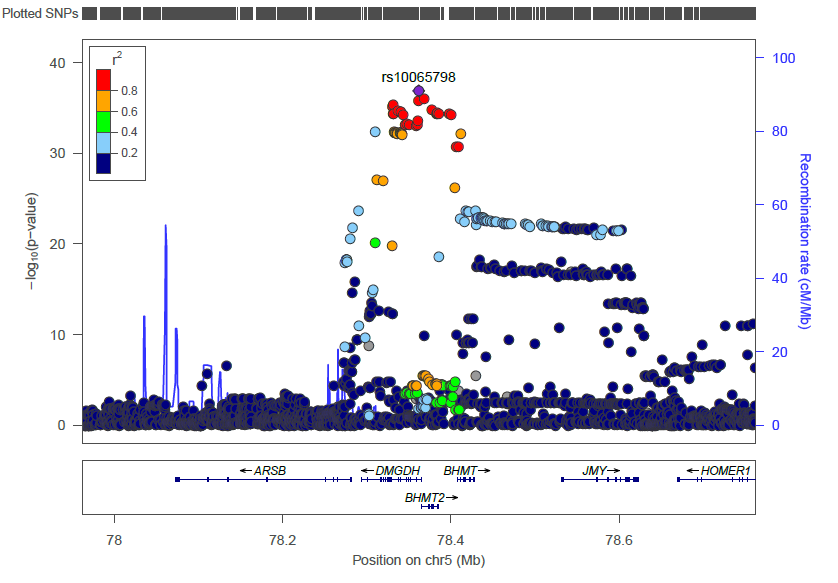

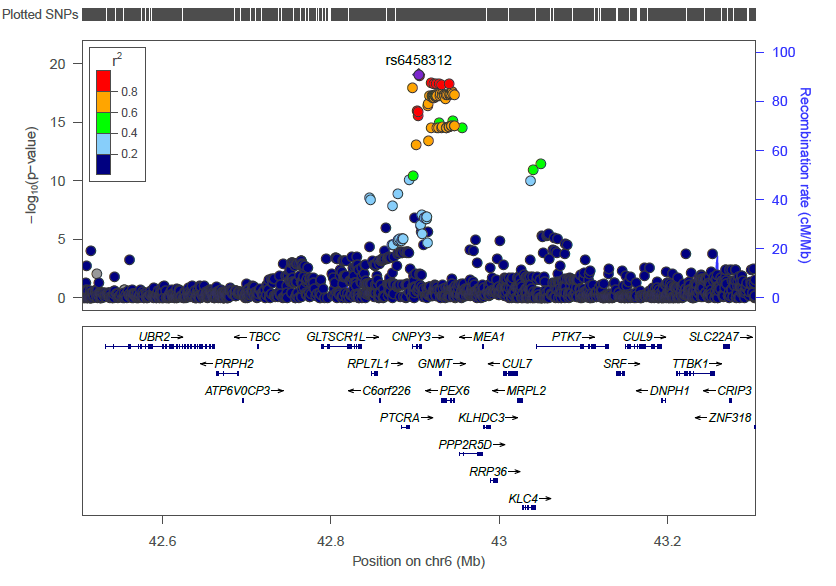

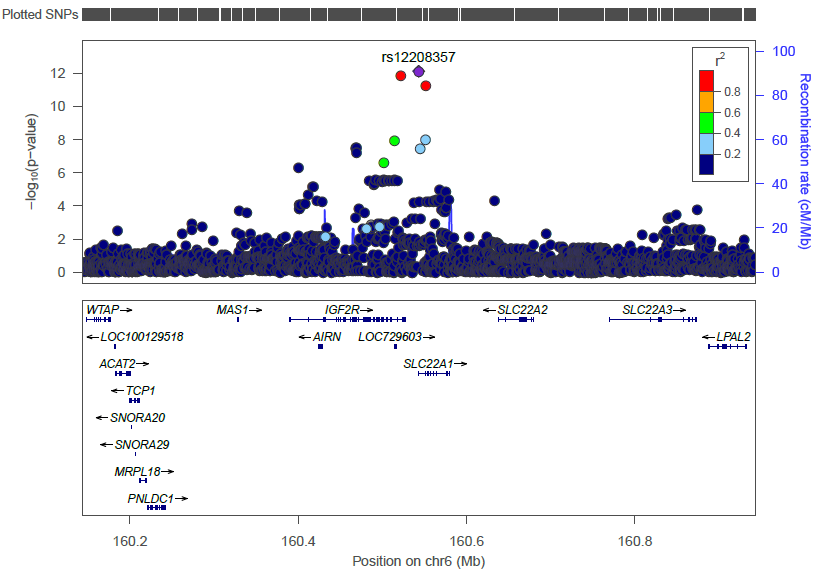

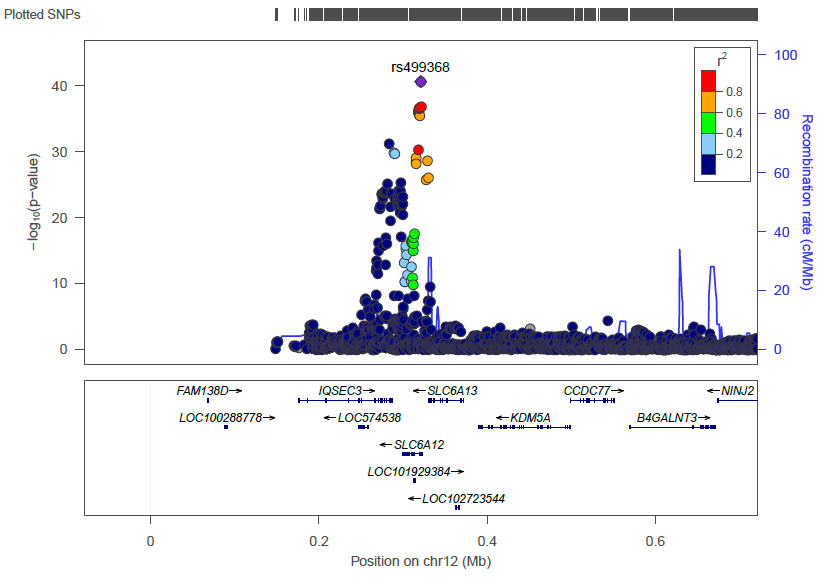

**Supplementary Figure 3.** Regional plots of lead genetic variants associated with circulating metabolite levels (1 of 3).

C. **Carnitine (3)**

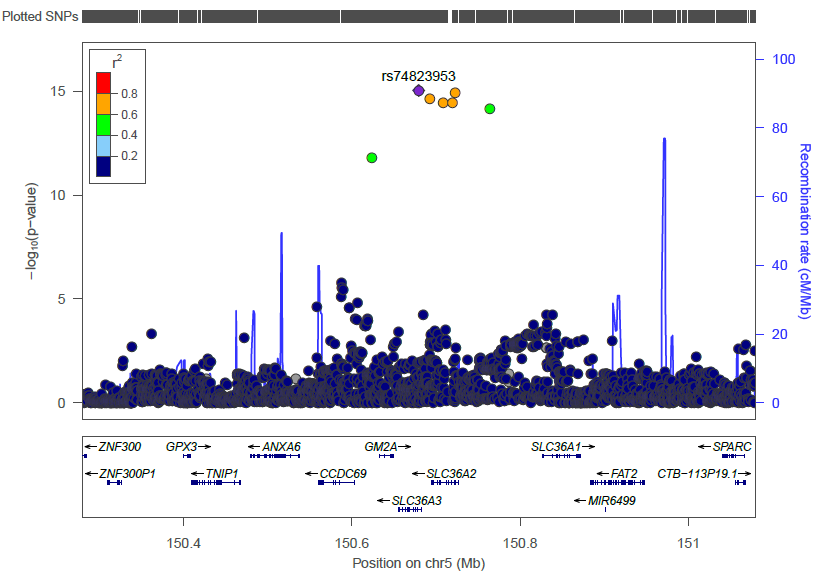

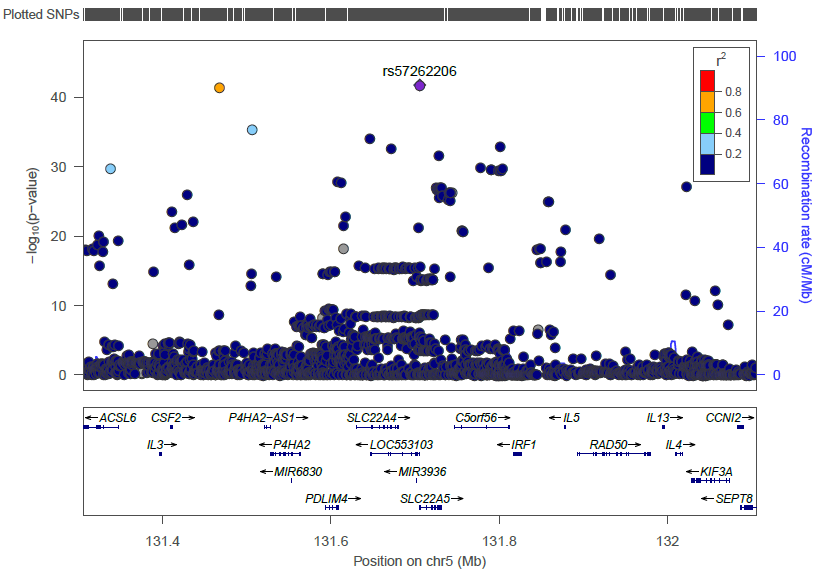

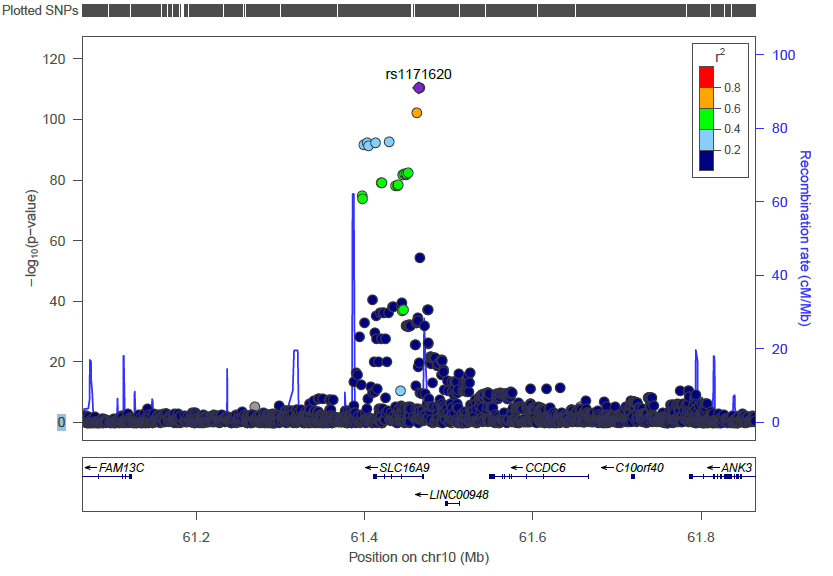

**Supplementary Figure 3.** Regional plots of lead genetic variants associated with circulating metabolite levels (2 of 3).

D. **Deoxycarnitine (3)**

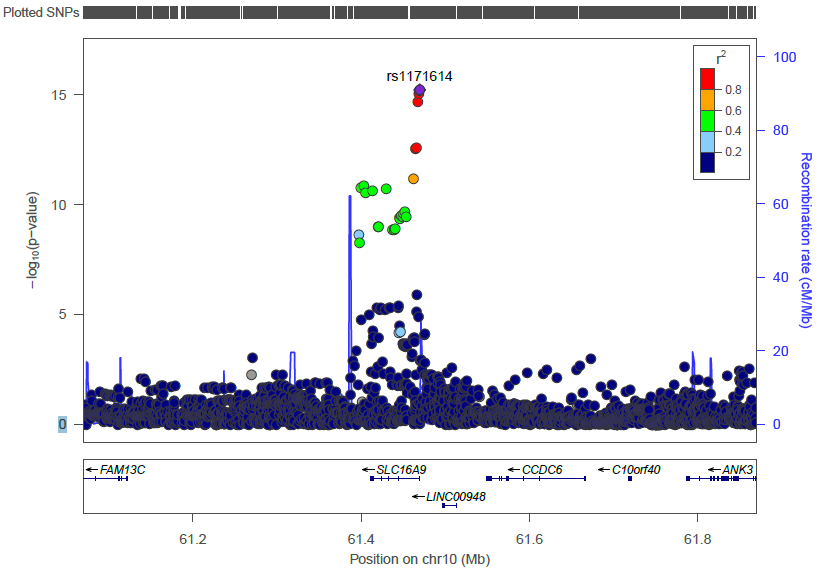

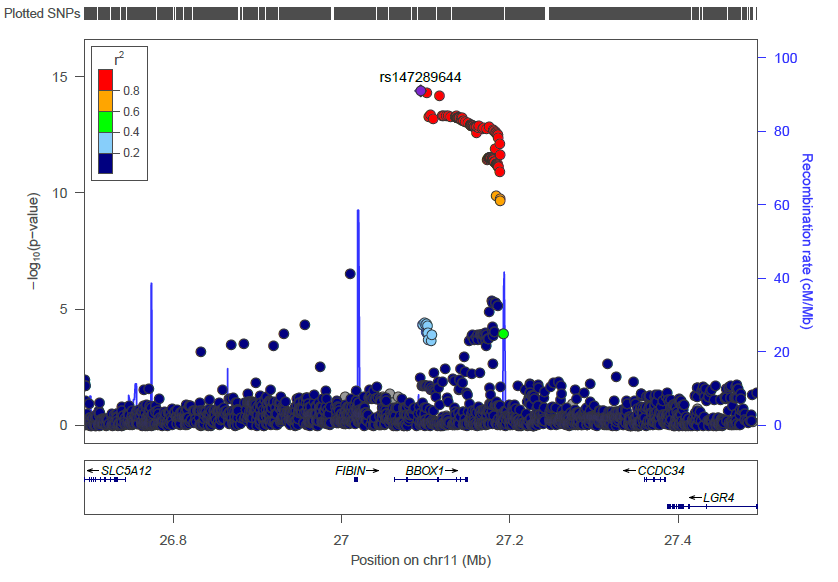

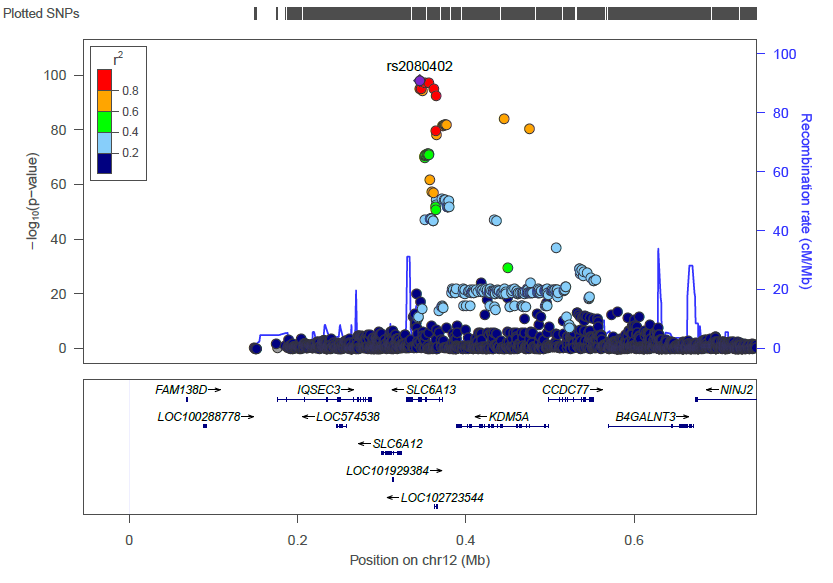

E. **Choline (3)**

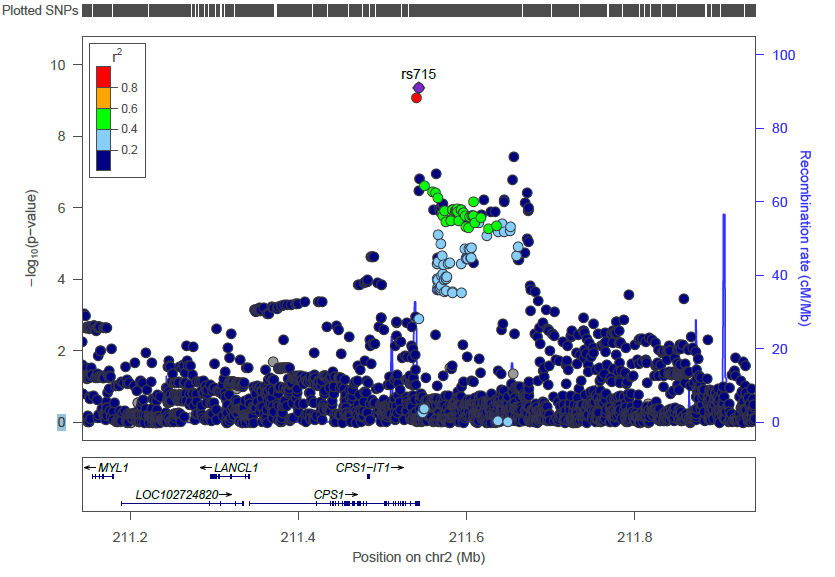

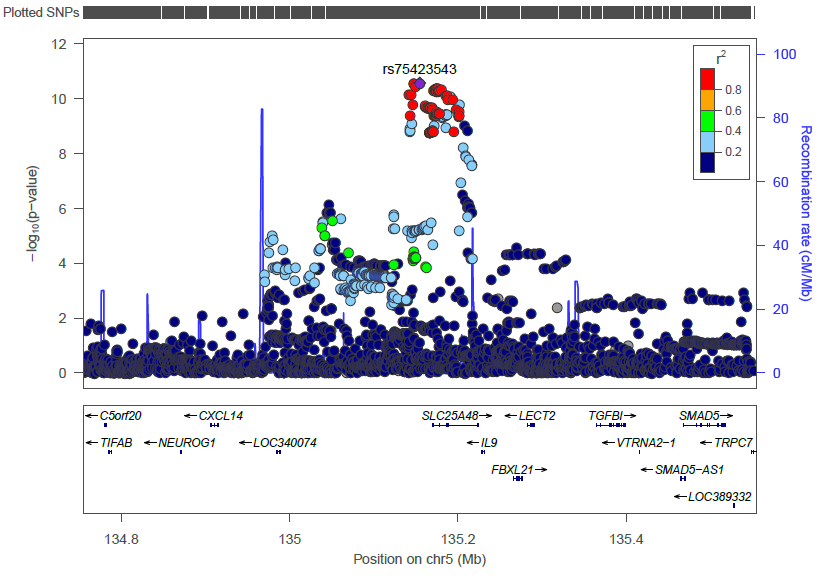

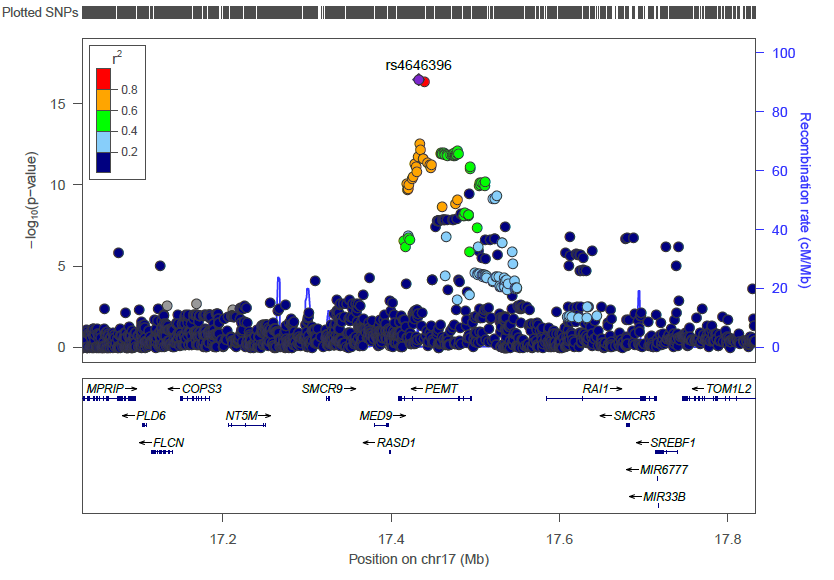

**Supplementary Figure 3.** Regional plots of lead genetic variants associated with circulating metabolite levels (3 of 3).

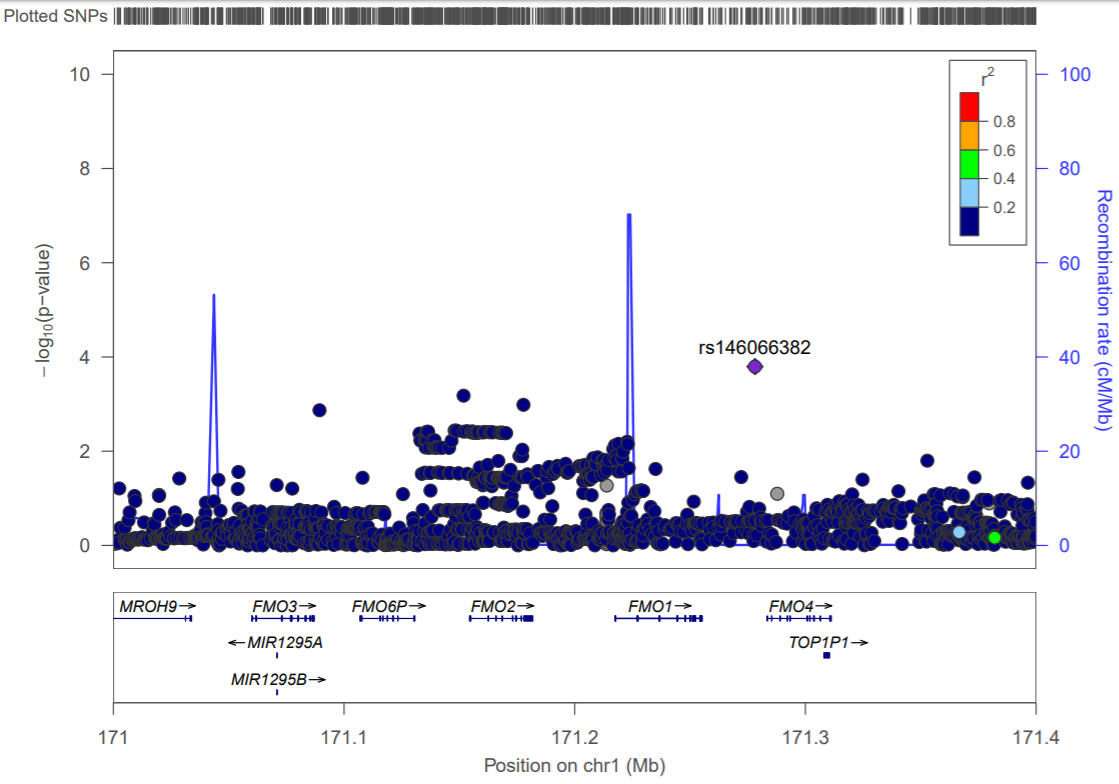

**Supplementary Figure 4.** Regional plot for the region encompassing FMO genes based on the GWAS summary statistics for TMAO.

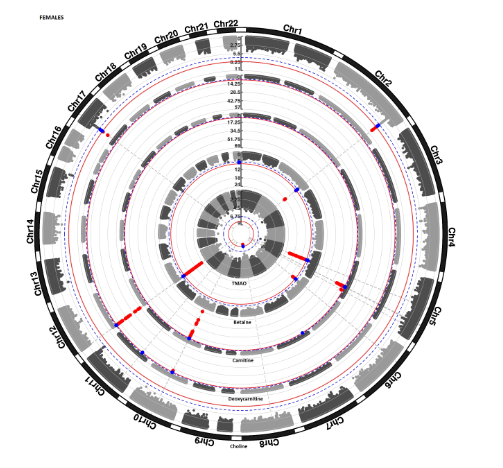

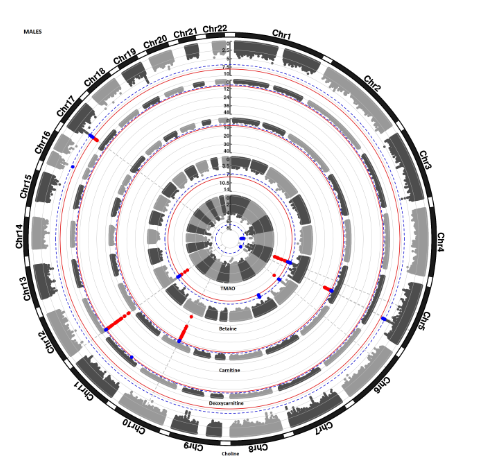

**Supplementary Figure 5.** Results of sex-stratified genome-wide association analysis for individual metabolites in females (first panel) and males (second panel). Each dot represent a genetic variant. Genetic variants surpassing Bonferroni corrected significance threshold (*p*-value < 8.33×10^-9)^ are highlighted in red. Genetic variants showing suggestive evidence of association (*p*-value < 1.7×10^-7^) are highlighted in blue.

|  | h2 |
| --- | --- |
| TMAO | 0.2061 |
| Betaine | 0.2765 |
| Carnitine | 0.178 |
| Deoxycarnitine | 0.1624 |
| Choline | 0.16 |
| TMAO_betaine | 0.1959 |
| TMAO_carnitine | 0.1739 |
| TMAO_deoxycarnitine | 0.1739 |
| TMAO_choline | 0.1436 |

**Supplementary Figure 6.** SNP-based heritability estimated using LD-score regression**.** Metabolite names are listed along *x*-axis, while heritability estimates are shown on *y*-axis.

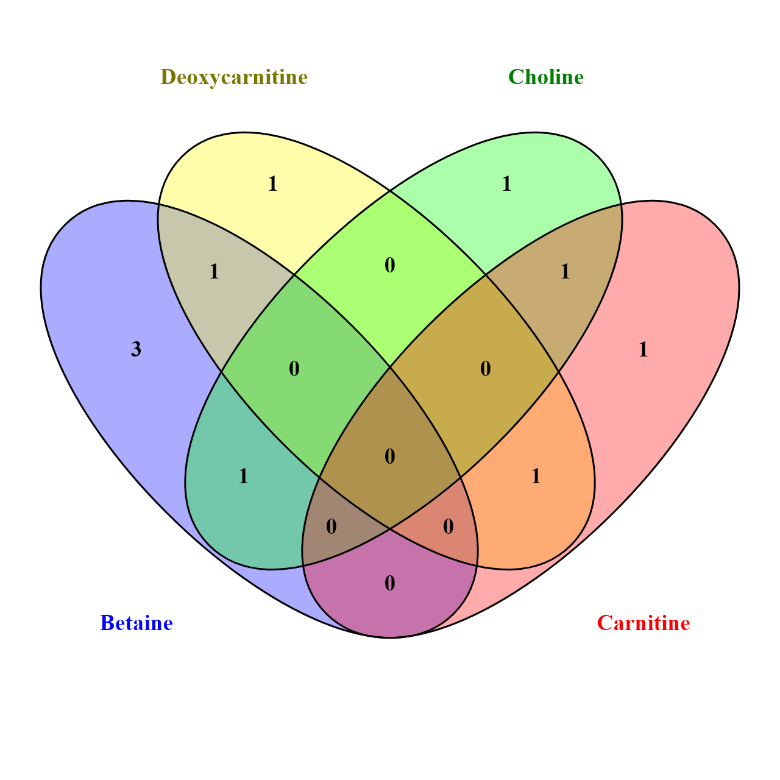

**Supplementary Figure 7.** Overlap of lead genomic loci identified in the GWAS of TMAO and its precursors.

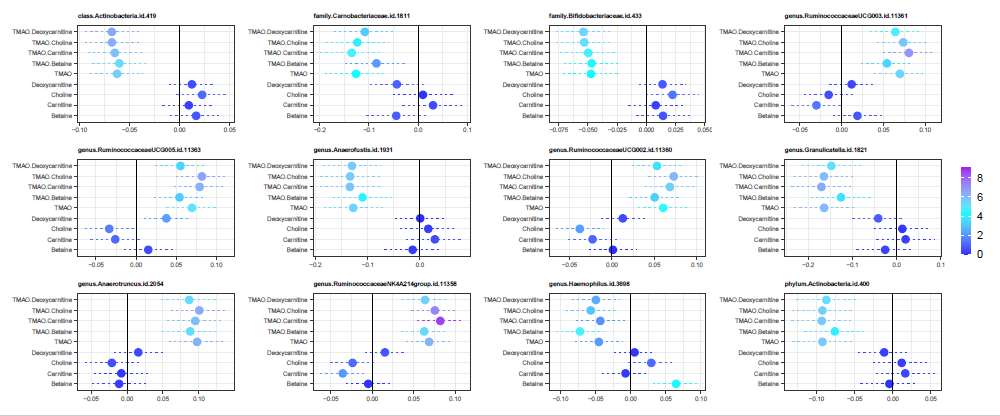

**Supplementary Figure 8.** Top associations of microbial taxonomy with metabolites. Each panel shows the estimated effect size (*x*-axis) of the association between metabolite abundance (*y*-axis) and a taxonomy abundance (panel title). Dashed lines display 95% confidence intervals for the estimates. Vertical lines highlight 0 effect. Color represents -log10(*p*-value).

A. Males	B. Females
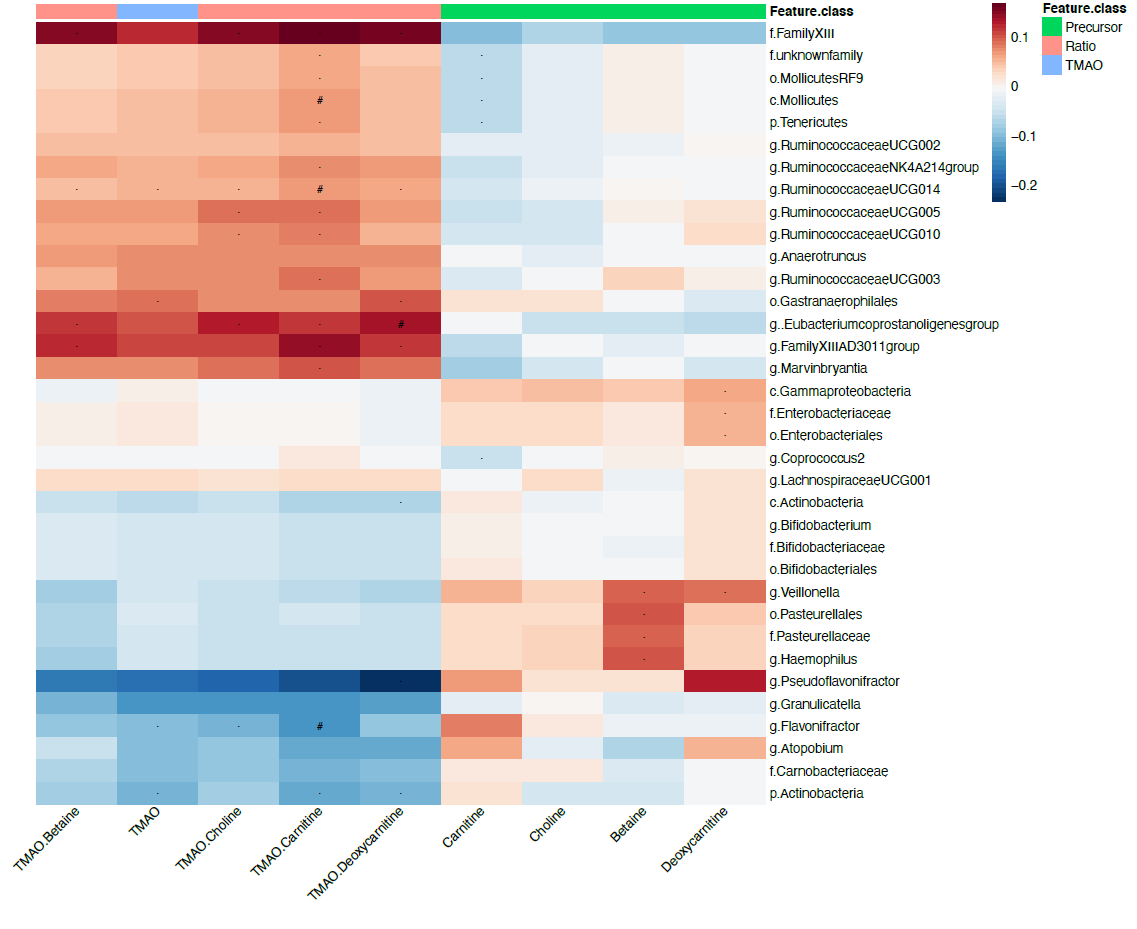

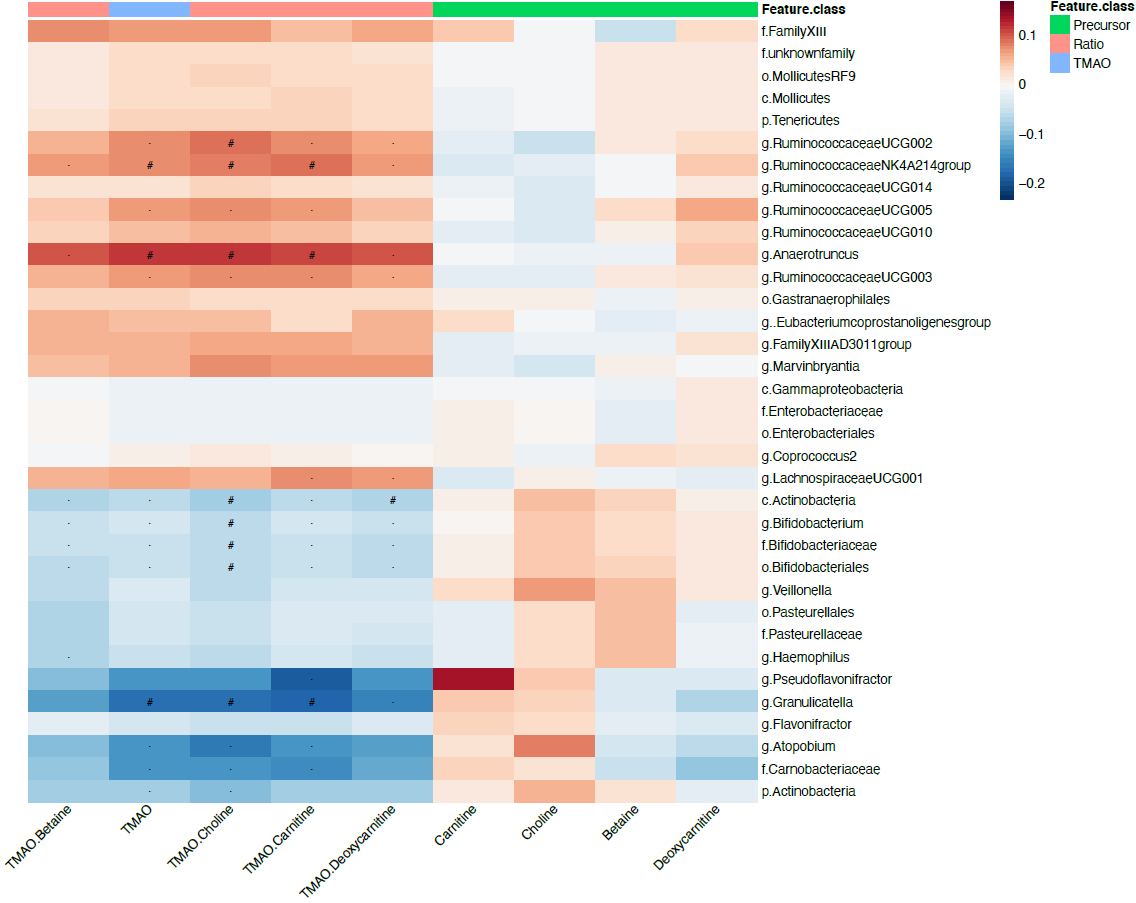

**Supplementary Figure 9.** Results of association analysis between metabolites and gut microbial taxa in males (a) and females (b). Metabolites are displayed on *x*-axis, and the microbial taxa on *y*-axis. Red color stands for positive correlation while blue color denoted negative correlation. The associations that surpassed Bonferroni corrected threshold for multiple testing (*p-*value < 6.2×10^-5^) are denoted by hash symbol, while star denotes suggestive associations with *p-*value < 1.2×10^-3^.

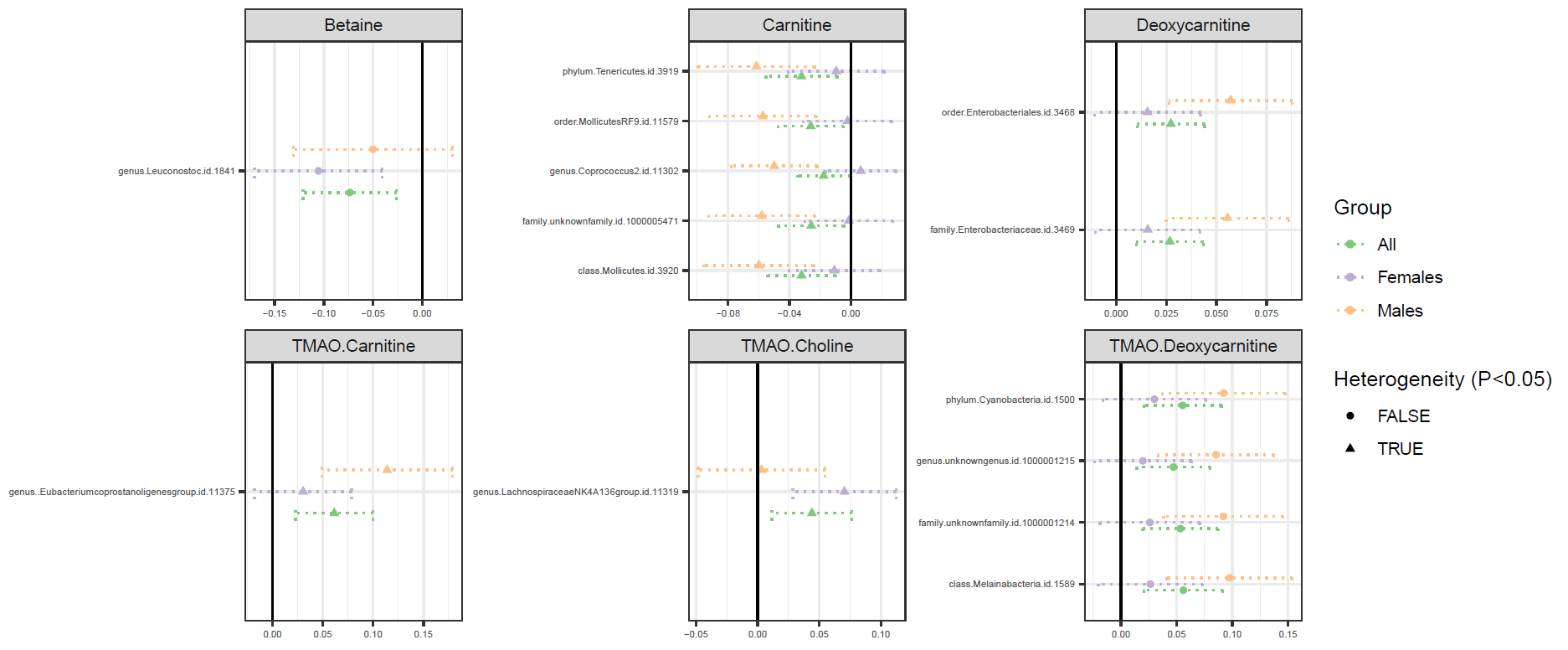

**Supplementary Figure 10.** Gender-heterogeneous associations of microbial taxonomy with metabolites. Each panel shows the estimated effect size (*x*-axis) of the association between bacterial taxa abundance (*y*-axis) and a metabolite (panel title). Point’s shape represents whether heterogeneity statistic Cochran's Q was under a *p*-value of 0.05. Dashed lines display 95% confidence intervals for the estimates. Vertical lines highlight 0 effect.

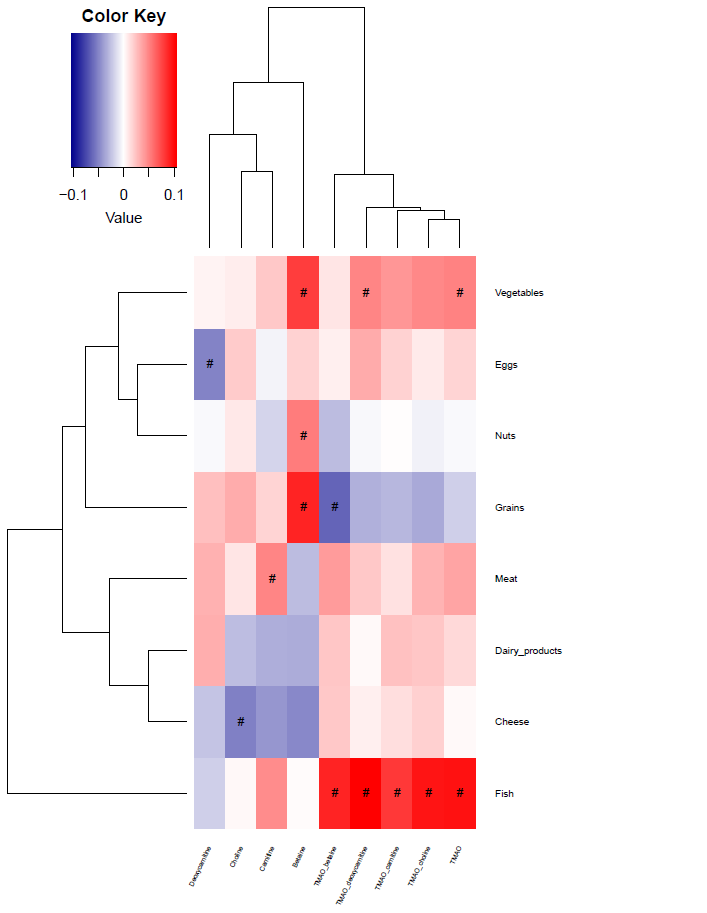

**Supplementary Figure 11.** Results of correlation analysis between metabolites and ratios and food groups. Metabolites are displayed on *x*-axis, and the food groups on *y*-axis. Red color stands for positive correlation while blue color denoted negative correlation. The associations that surpassed Bonferroni corrected threshold for multiple testing (*p*-value < 7.58 × 10^-4^) are denoted by hash symbol.
